# A Biopsychosocial Risk Score for Stratifying Disease Vulnerability in Healthy Populations: A Prospective Cohort and Multi-Omics Study in the UK Biobank

**DOI:** 10.64898/2026.02.08.26345832

**Authors:** Jin Chen, Congying Chu, Miguel Garcia-Argibay, Wei Li, Christos Christogiannis, Tianye Jia, Charlotte Walton, Sangma Xie, Tifei Yuan, Samuele Cortese, Bing Liu, Jiaojian Wang

## Abstract

Proactive identification of systemic vulnerability for disease(s) before clinical onset in healthy individuals is an ultimate goal of preventive and precision medicine, yet current tools remain largely disease-specific and fail to quantify latent vulnerability, an integrative measure of underlying health status, for early prevention and risk-stratified intervention. To address this, we developed the Risk Score for Disease Vulnerability (RS4DV) based on 85 accessible biopsychosocial measures, which was constructed using a Light Gradient Boosting Machine trained and validated in the UK Biobank (*n* = 391,193). Its capacity to capture pre-clinical vulnerability was subsequently evaluated in a held-out cohort free of baseline diagnoses (*n* = 35,193). Over a median follow-up of 14.7 years, baseline RS4DV stratified long-term health outcomes in the held-out cohort, in which high-risk individuals exhibited accelerated disease accumulation (HR = 2.53, 95% CI: 2.44-2.62) and elevated risk of all-cause mortality (HR = 4.03, 95% CI: 3.68-4.41). Multi-omics analyses further revealed that RS4DV captures signatures of systemic inflammation, metabolic dysregulation, and accelerated brain ageing, establishing its biological interpretability. To facilitate real-world translation, we developed a practically feasible version of the RS4DV with only six routinely accessible items, which maintained robust predictive fidelity relative to the full model. This light-version model demonstrated robust generalizability in the 1970 British Birth Cohort under a zero-shot paradigm. Collectively, RS4DV provides a biologically grounded and scalable tool for personalized risk assessment decades before clinical onset and early-stage risk stratification, enabling a paradigm shift toward proactive health management and precision prevention.

## Introduction

Population ageing and ongoing epidemiological transition have fundamentally reshaped the global burden of disease, with chronic and non-communicable conditions now dominating morbidity and mortality^1–3^. As life expectancy increases, a growing proportion of individuals live with multiple chronic conditions simultaneously, making multimorbidity the norm in modern healthcare systems^4,5^. Although medical specialization has driven remarkable advances in diagnosis and treatment^6,7^, its disease-centric framework remains inadequately aligned with the complex and overlapping nature of multimorbidity in ageing populations. A critical unmet need is the ability to efficiently and accurately stratify ostensibly healthy individuals according to their risk of progressing to overt multimorbidity, a quantitative foundation essential for advancing primordial prevention. Crucially, we posit that the burden of multimorbidity is an observable manifestation of individuals’ integrative health status. Consistent with the syndemics framework^8,9^, diseases rarely occur in isolation but co-emerge through shared biological pathways such as chronic inflammation^10^, immune dysregulation and metabolic dysfunction^11^, together with common social-psychological drivers like socioeconomic disadvantage, chronic stress and proinflammatory phenotype^12,13^. This recognition reemphasizes the need for a quantitative indicator of an individual’s integrative health status, which is across conventional organ-based boundaries and captures an individual’s vulnerability to the co-occurrence of chronic and non-communicable diseases.

Notably, this integrative view of health has long been articulated in the biopsychosocial (BPS) model^14^, which conceptualizes disease as arising from the dynamic interplay of biological, psychological, and social processes^15^. However, its translation into clinical practice and empirical research has historically been constrained by limited data availability and methodological capacity. The emergence of large-scale population cohorts (e.g., UK Biobank), extensive electronic health records, and advanced machine-learning techniques creates an unprecedented opportunity to revisit and operationally realize the BPS model. Moreover, machine-learning approaches are particularly suited to integrate multidimensional data such as biological indicators (e.g., blood pressure), psychometric constructs (e.g., neuroticism scores) and social determinants (e.g., socioeconomic position) to capture the high-dimensional and non-linear interactions related to multimorbidity^16–18^. Therefore, quantifying the vulnerability to multimorbidity can be conceptualized as an emergent property of the integrated BPS system.

Furthermore, the accessibility and cost-effectiveness of BPS measures in clinical settings make them ideal instruments for population-wide health surveillance. Despite this advantage, there exists a critical gap lacking an established modelling framework capable of mapping these upstream BPS determinants to an individual’s latent vulnerability to multimorbidity. Additionally, it remains unproven whether such a BPS-informed model can achieve robust generalizability across independent cohorts while providing biologically meaningful interpretability beyond predictive accuracy alone. Addressing these challenges requires a risk-scoring model having generalizability^19^ and biological interpretability^20,21^. In terms of generalizability, rigorous validation across large population-based cohorts through extensive out-of-sample testing is essential^22^. Regarding biological interpretability, it demands that disease vulnerability evaluated by the model connects to underlying biological mechanisms, such as molecular^23^, genetic^24^, and neural factors^25^.

Therefore, in this study, we developed the Risk Score for Disease Vulnerability (RS4DV), a machine learning-based metric integrating a range of biological, psychological, and social determinants of health. RS4DV aims to quantify the risk of health deterioration at the preclinical stage, capturing how BPS determinants collectively shape vulnerability to multimorbidity. Specifically, we constructed the RS4DV using the UK Biobank cohort and subsequently evaluated it in the held-out test samples (Figure 1.I). (Figure 1.I). Its applicability at the preclinical stage was then demonstrated in its ability to stratify longitudinal disease accumulation and all-cause mortality (Figure 1.II). To delineate the biological interpretability of RS4DV, we further conducted multi-omics analyses, including genome-wide association studies, proteomics, and neuroimaging (Figure 1.III). Finally, to facilitate real-world translation, we derived and validated a light version of RS4DV that retained performance comparable to the full model. This simplified model generalized well to an independent cohort (the 1970 British Birth Cohort) under a zero-shot paradigm and could be further efficiently fine-tuned (Figure 1.IV). Together, these analyses establish RS4DV as a clinically feasible and biologically grounded framework for quantifying disease vulnerability across populations, bridging the gap between upstream BPS determinants and downstream risk of health deterioration at the preclinical stage.

**Figure 1.**
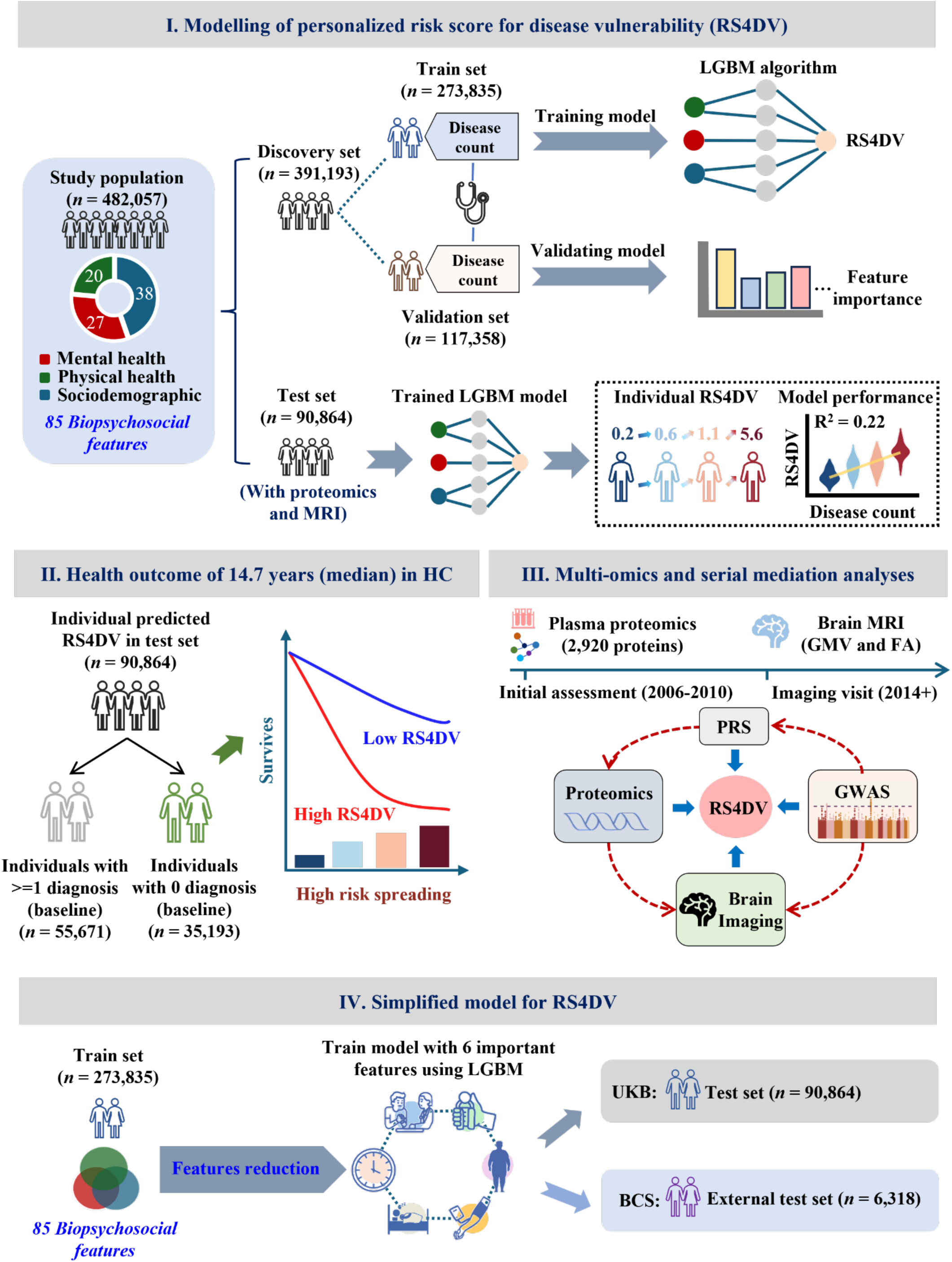
Schematic overview of the study design and analytical framework for RS4DV. (I) RS4DV was developed in the UK Biobank cohort (*n* = 482,057) based on 85 BPS features spanning mental health, physical health, and sociodemographic domains. Participants were split into a discovery set (*n* = 391,193), which was further randomly divided into a training set (*n* = 273,835) and a validation set (*n* = 117,358), and an independent test set (*n* = 90,864, with proteomic and MRI data available). A Light Gradient Boosting Machine (LGBM) model was trained to predict disease vulnerability, defined as the cumulative number of diagnoses before baseline. Feature importance was evaluated in validation set, model performance and individualized RS4DV values were derived in test set. (II) The prognostic value of RS4DV for risk stratification of future disease was assessed in the test set over a median follow-up of 14.7 years. Participants free of disease at baseline (individuals with 0 diagnoses; *n* = 35,193) were stratified by RS4DV to examine whether higher baseline RS4DV was associated with increased risk of incident disease and higher risk of all-cause mortality. (III) To investigate biological underpinnings, we performed multi-omics and serial mediation analyses in the test set. Plasma proteomics and brain MRI measures (grey matter volume, GMV, and fractional anisotropy, FA) were integrated with polygenic risk scores (PRS). RS4DV was positioned as a mediator linking genetic risk, proteomic signatures, and brain imaging correlates. (IV) Finally, a simplified RS4DV model was derived by reducing the feature set to 6 key predictors using LGBM. This light-version model was validated in the UK Biobank test set (*n* = 90,864) and independently replicated in 1970 British birth cohort (BCS70; *n* = 6,318).

## Results

### Cumulative disease burden as a quantitative phenotype of systemic vulnerability

Systemic vulnerability of multimorbidity represents a latent health state reflecting the cumulative impact of biological, psychological, and social stressors. As such, it cannot be directly observed. We conceptualized the accumulation of chronic and non-communicable diseases as an observable manifestation of this latent vulnerability, analogous to deficit accumulation in frailty research^26,27^. Each additional diagnosis reflects a further loss of systemic resilience, providing a quantitative and clinically meaningful proxy of underlying health status. Thereby in the UK Biobank cohort, the total number of recorded diagnoses within a predefined disease spectrum was used as an index of cumulative disease burden and served as the outcome variable for training the RS4DV model. This disease spectrum was defined in our previous studies^28–30^ through a systematic review and primarily included major chronic and non-communicable conditions that substantially influence on overall health status (Supplementary Table S1).

Following the Preprocessing of missing biopsychosocial features (see Methods for details), a total of 482,057 participants in the UK Biobank cohort were included in the present analyses (220,509 males, 45.7%; 261,548 females, 54.3%). At the time of enrolment (baseline) 28% of the cohort had accumulated two or more diseases (*n* ≥ 2) until the enrolment by UK Biobank. To overview multimorbidity patterns, the included diseases were grouped by ICD-10 chapters, and joint probabilities across chapters were calculated (Supplementary Figure S1A). We found that the coexistence of diseases and symptoms was not confined to a single system but extended across different health domains. For example, individuals with mental health conditions were often also diagnosed with circulatory diseases (Joint Probability = 3.6%), a phenomenon observed across other major diagnostic groups (Supplementary Figure S1A-B). To validate the phenotypic relevance of our cumulative disease count, we examined its association with self-rated overall health (SRH) scores (Data-Field 2178), which is a well-established proxy for general physiological status and a robust predictor of mortality^31^. We observed a stepwise degradation in SRH score as the disease count increased (Supplementary Figure S2). This consistence confirms that our objective multimorbidity metric effectively captures the biological reality of health deterioration.

### Development and validation of the RS4DV model

The included 482,057 participants were further divided into a discovery cohort (*n* = 391,193) for model development and a held-out independent test cohort (*n* = 90,864) for external evaluation. The test cohort specifically included participants with proteomic or neuroimaging data to facilitate downstream mechanistic analyses. Key demographic and socioeconomic characteristics were well-balanced between the discovery and test cohorts (SMD < 0.1; Supplementary Table S4). During the training phase, the discovery cohort was randomly partitioned into a training set (*n* = 273,835) and an internal validation set (*n* = 117,358) at a 7:3 ratio. Within the training set, a Light Gradient Boosting Machine (LGBM) model was trained using a training-validation framework with hyperparameter optimization and early stopping in the internal validation set to control overfitting. The optimized model demonstrated robust and consistent predictive performance across datasets (training R² = 0.22, validation R² = 0.20, test R² = 0.18, Figure 2A). No substantial performance decay was observed between training and independent test cohorts, indicating effective control of overfitting and generalizability of the model. The trained model further exhibited high calibration in the test cohort (slope = 1.06, intercept = 0.03), with predicted risks closely aligned with observed disease burden.

**Figure 2.**
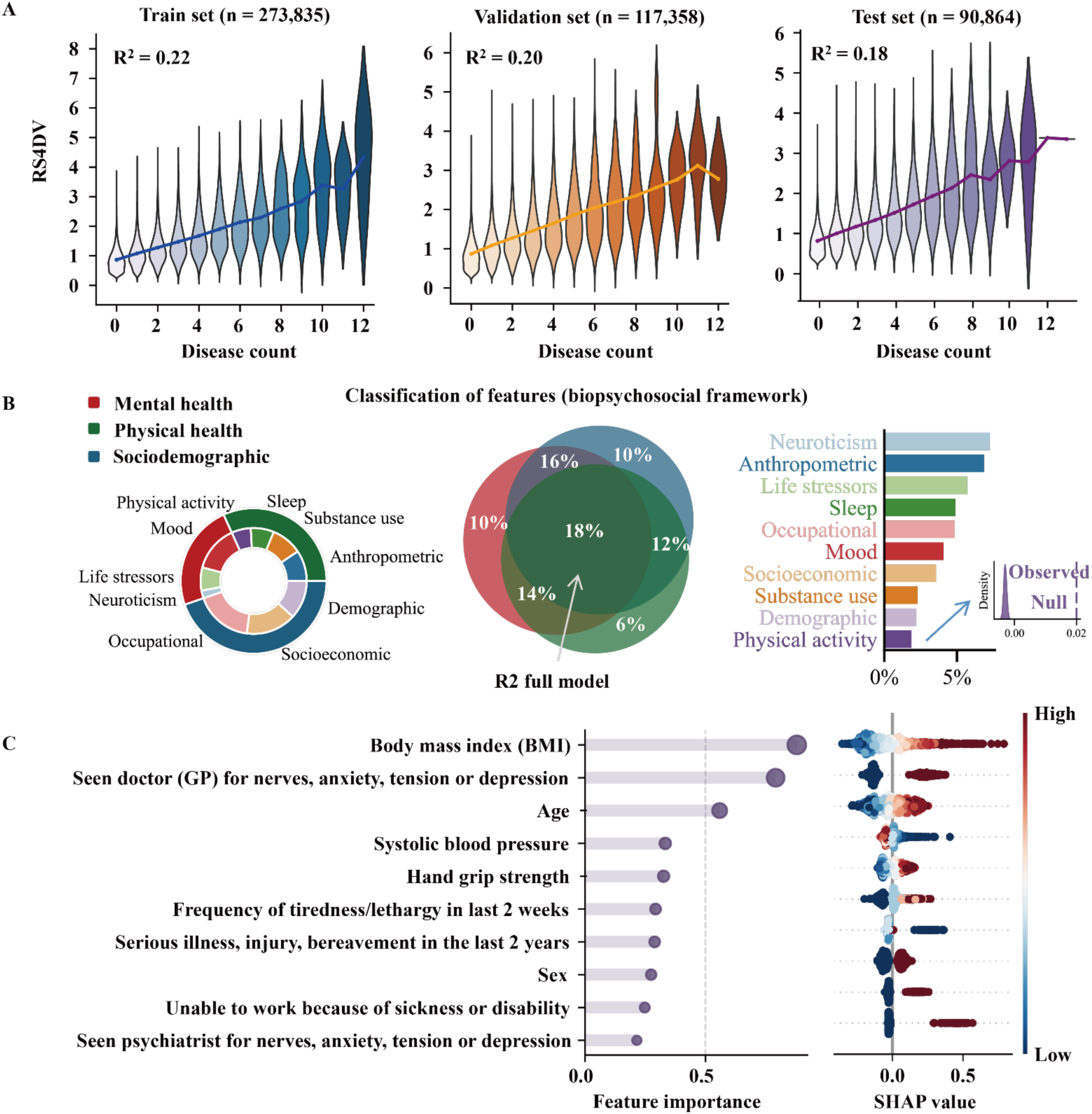
Multivariate modelling of disease vulnerability using a BPS framework. **(A)** LGBM model performance in the training (*n* = 273,835), validation (*n* = 117,358), and independent test (*n* = 90,864) sets, where the test set includes participants with proteomic and brain imaging data. Violin plots depict the relationship between predicted risk score for disease vulnerability (RS4DV) from the final LGBM model and the actual disease counts. **(B)** Schematic of the 85 candidate features encompassing physical, mental, and social domains, classified under a biopsychosocial (BPS) framework. Left: Donut chart illustrating the proportion of features within each domain. Middle: Pie chart showing the variance explained (R^2^) in the test set by models trained using features from each domain separately. Right: Bar chart presenting the variance explained (R^2^) in the test set by models trained on each of the 10 feature subsets. Statistical significance was assessed using permutation tests; the model trained on the lowest-performing subset (lowest R²) still demonstrated significant explanatory power. **(C)** Ranking of the top 10 features by mixed feature importance in the LGBM model, with their respective SHAP values indicating each feature’s contribution to the predicted disease counts. Positive SHAP values suggest a higher predicted disease vulnerability, whereas negative values indicate a protective or reducing effect.

We next evaluated the contribution of integrating multiple BPS domains in the independent test cohort. The model incorporating all three domains showed the strongest predictive performance (R² = 0.18), outperforming models based on single domains (R² = 0.06-0.10) or any two-domain combinations (R² = 0.12-0.16; Figure 2B). Moreover, by conducting model interpretation with the combination of Gain, Split and SHapley Additive exPlanations (SHAP) values^32^, we identified BMI, mental health history, age, systolic blood pressure, and grip strength as the top-5 dominant predictors in the trained model (Figure 2C; Supplementary Table S5).

Furthermore, the robustness of RS4DV was supported by multiple sensitivity analyses. To evaluate the potential influence of missing data imputation on model estimates, we conducted a sensitivity analysis based on complete-case samples. Specifically, a subset of participants with complete measurements across all required BPS features (*n* = 20,248) was extracted from the discovery cohort and used to retrain the model without imputation. In the independent test cohort, RS4DV scores derived from the complete-case model showed high consistency with those from the primary imputed model (*ρ* = 0.95, *p* < 0.001; Supplementary Figure S5). To assess the dependence of RS4DV on disease definition, we conducted a sensitivity analysis within the UKB (Category 1712) by restricting outcome construction to ICD-10 conditions with prevalence exceeding 1% and 5%, respectively (Supplementary Table S6, S7). The model retrained using these high-prevalence disease sets generated RS4DV scores that remained highly consistent with those from the primary model in the independent test cohort (*ρ* = 0.90/0.95, *p* < 0.001; Supplementary Figure S6). In addition, to evaluate the sex effect on RS4DV, we developed two sex-specific models. Both models showed highly consistent predictive performance with the primary model (*ρ* = 0.99/0.99, *p* < 0.001, Supplementary Figures S7–S8).

### Longitudinal stratification value of RS4DV for multimorbidity and mortality in the baseline disease-free population

We evaluated the prospective performance of RS4DV in participants free of any diagnosed disease at recruitment (*n* = 46,550 in the internal validation set; *n* = 35,193 in the held-out test set). Over a median follow-up of 14.7 years, baseline RS4DV demonstrated robust long-term predictive ability. Higher RS4DV values were significantly associated with more disease accumulation in both subsets (validation subset, *ρ* = 0.32, *p* < 0.001, Figure 3A; test subset, *ρ* = 0.30, *p* < 0.001, Figure 3B). Furthermore, Kaplan-Meier analyses revealed significant risk stratification, demonstrating RS4DV to be an effective predictor of future health decline. Individuals (no disease diagnosis at the baseline) in the top decile of RS4DV experienced faster disease accumulation and reduced survival compared with those in the bottom decile (10% decile: validation set: *p* < 1×10^-300^, test set: *p* = 4.97×10^-281^; 30% decile: validation set: *p* < 1×10^-300^, test set: *p* < 1×10^-300^; Figure 3C&D). By year 15, the disease accumulation in the high-risk group reached 64.5% (vs. 18.4% in low-risk group) in the validation set, with similar gaps observed in the test set (62.9% vs. 21.5%; Figure 3C). Mortality trends were also significantly distinct (Figure 3D), with cumulative rates of 13.3% vs. 1.6% (validation set) and 11.2% vs. 1.0% (test set). These findings were then supported by Cox proportional hazards models. When modelled as a continuous variable, RS4DV was strongly associated with incident multimorbidity (validation set Hazard ratio (HR) = 2.57, test set HR = 2.53, both *p* < 1×10^-300^) and all-cause mortality (validation set HR = 2.99, test set HR = 4.03, both *p* < 1×10^-198^). Disease-specific analyses further demonstrated that higher RS4DV consistently predicted elevated risks across a broad range of diseases. Participants in the top decile exhibited significantly increased incidence for 25 diseases compared with the bottom decile, and similar patterns were observed for the top 30% versus bottom 30% groups (Supplementary Figures S9-S10). High-risk individuals showed a predominance of cardiometabolic and mental health disorders, whereas incident conditions in low-risk individuals were more randomly distributed (Supplementary Figure S11).

**Figure 3.**
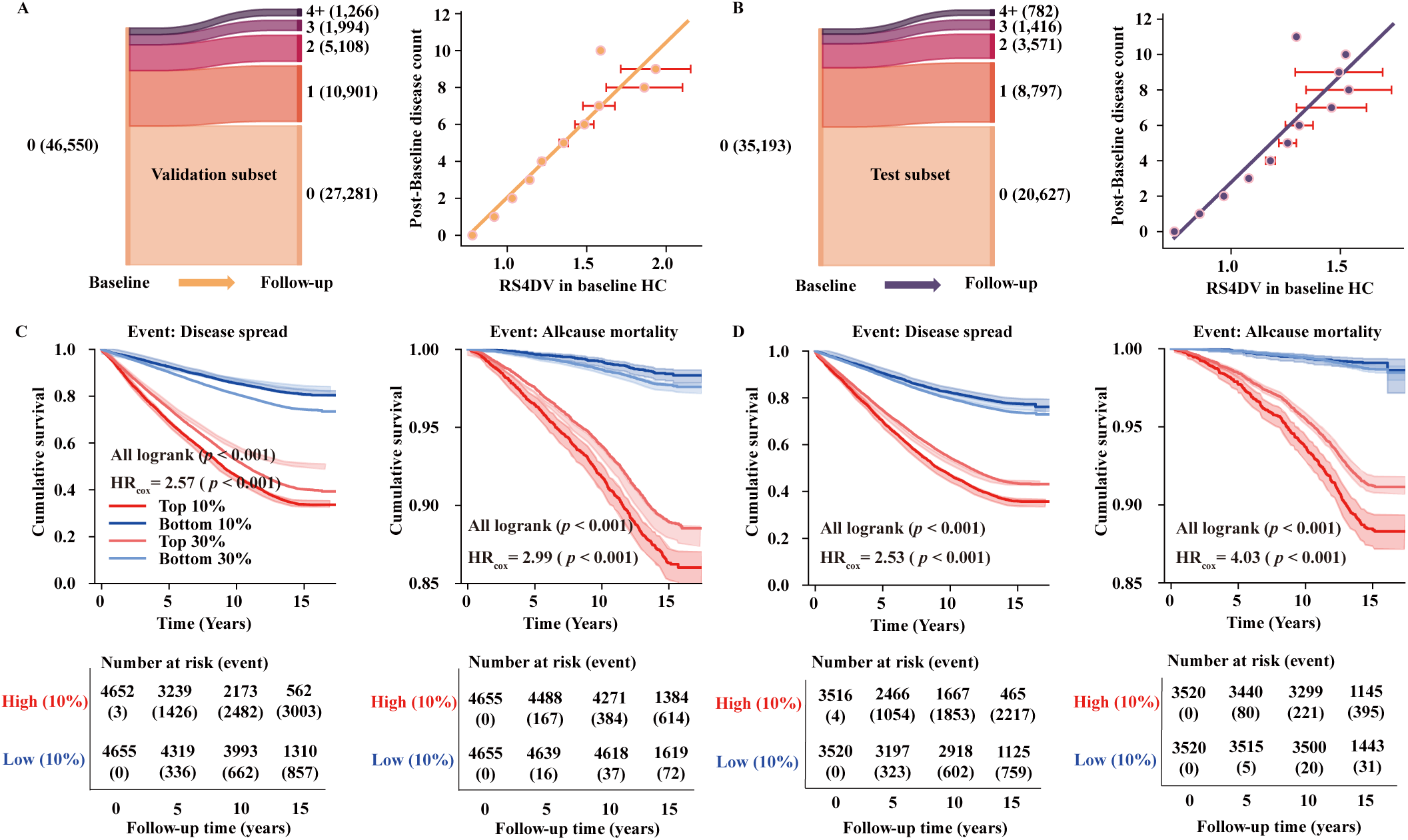
RS4DV-based longitudinal assessment in disease-free participants. **(A)** In the validation subset (*n* = 46,550), a Sankey diagram (left) depicts transitions of disease status from baseline to follow-up in individuals who were disease-free at baseline. Each flow’s width corresponds to the proportion of participants transferring from one disease category at baseline (e.g., 0, 1, 2, 3, or 4+ diseases) to a specific number of disease categories at follow-up. On the right, a scatter plot shows the relationship between the baseline RS4DV (x-axis) and the total number of diseases recorded at follow-up (y-axis). **(B)** In the independent test subset (*n* = 35,193), the same approach is used to visualize the progression from baseline disease-free status to follow-up number of disease categories (left) and to illustrate how the baseline RS4DV relates to the subsequent disease counts (right). **(C, D)** Kaplan-Meier survival curves illustrate the time-to-event analysis for participants classified into the top 10%, 30% (High Risk) and bottom 10%, 30% (Low Risk) of baseline RS4DV. Plots are shown for both the validation subset (left) and test subset (right) over average median follow-up time 14.7 years of follow-up. The x-axis indicates time since baseline (years), while the y-axis displays the cumulative survival probability. Numbers at risk (event) are provided below each curve to indicate how many individuals remain in the analysis at each time point. Hazard ratios (HRs) derived from univariable Cox proportional hazards models using RS4DV are displayed within the plots.

### Molecular and neurobiological underpinnings of RS4DV

We investigated the biological basis of RS4DV using integrative analyses across genomics, proteomics, and neuroimaging. A genome-wide association study (GWAS) in the individuals without baseline disease diagnosis (*n* = 127,898) identified 10 genome-wide significant loci (26 mapped genes, Figure 4A; Supplementary Table S8, S9). Although modest genomic inflation was observed (*λ* = 1.25, Supplementary Figure S12), LD score regression indicated that this reflected polygenicity rather than population stratification (Intercept = 1.03). Mapped genes were enriched in pathways related to MAPK cascade regulation, DNA repair, and intracellular protein transport, highlighting molecular processes relevant to inflammation, cell proliferation, and core cellular activities (Supplementary Figure 13A). The GWAS Catalog further indicated that these mapped genes were associated with various physical traits, such as grip strength and BMI and lifestyle factors including sleep patterns (Supplementary Figure 13B). To evaluate the downstream relevance and robustness of the GWAS findings, we constructed a polygenic risk score (PRS) based on the GWAS’s statistics and examined its performance in the held-out test cohort of individuals with at least one disease at baseline (*n* = 38,576). This cross-population validation strategy was adopted to minimize potential reverse causation in the discovery phase and to assess the generalizability of genetic signals in clinical populations. In the test cohort, the PRS was positively associated with baseline RS4DV (*ρ* = 0.06, *p* < 0.001; Supplementary Figure 14A, left) and was related to subsequent disease accumulation over a median follow-up of 14.7 years, with higher scores observed among individuals who developed multiple new conditions (Supplementary Figure S14A, right). Furthermore, RS4DV showed significant genetic correlations with cardiometabolic traits (obesity, diabetes) and psychiatric disorders (depression, anxiety), confirming that the score captures a shared genetic liability spanning physical and mental health domains (Supplementary Figure 14B).

**Figure 4.**
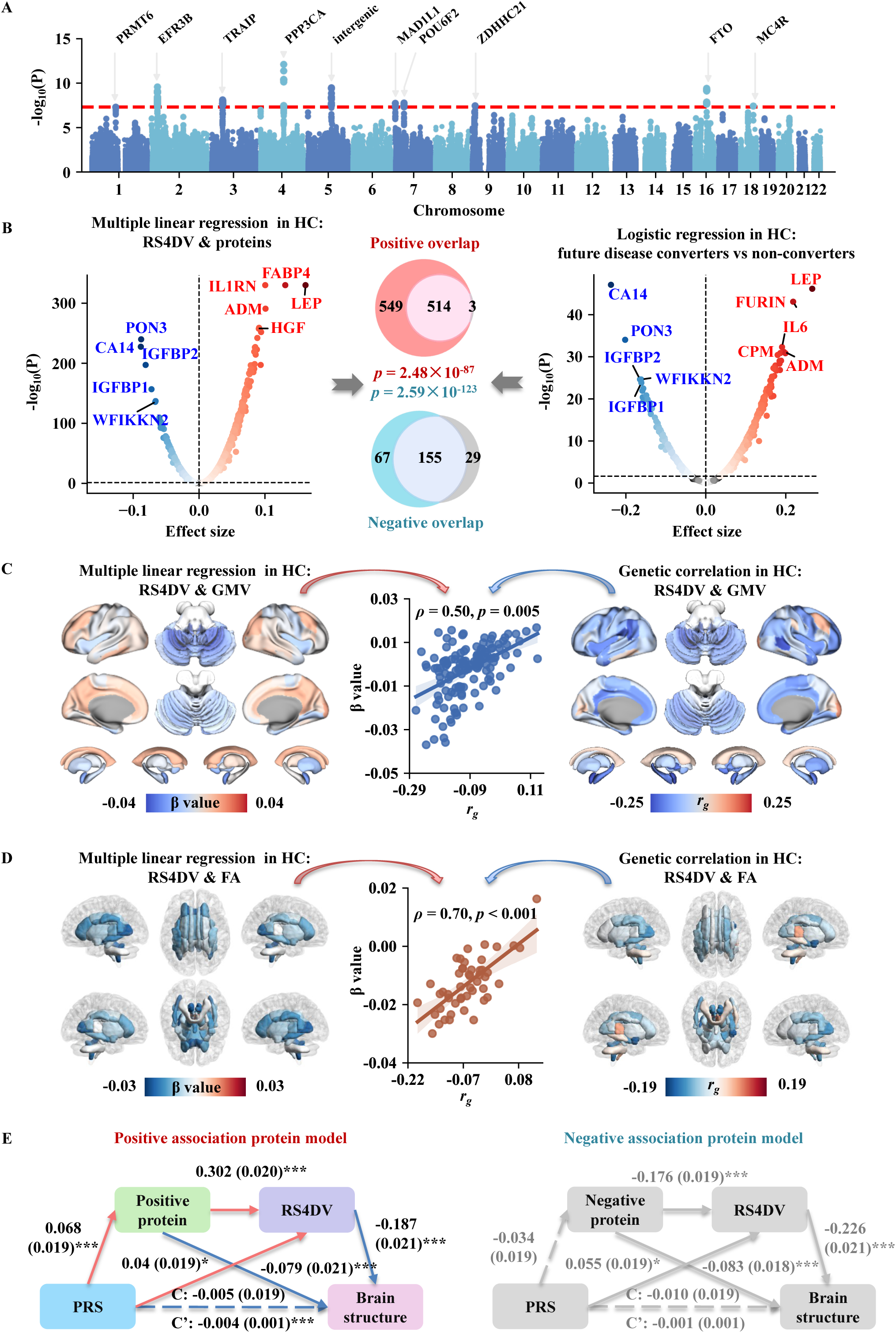
Genetic, proteomic, and neuroimaging correlates of RS4DV. **(A)** Manhattan plot of the genome-wide association study (GWAS) of RS4DV. The horizontal dashed line denotes the genome-wide significance threshold (*p* = 5 × 10⁻⁸). **(B)** Left: Volcano plot showing associations between baseline RS4DV and protein levels in disease-free participants based on multiple linear regression. Right: Volcano plot showing associations from logistic regression comparing participants who developed diseases during follow-up (coded as “1”) versus those who remained disease-free at both baseline and follow-up (coded as “0”). Effect sizes were estimated using two-sided tests, and proteins surpassing the Bonferroni threshold (*p* < 0.001) are annotated as upregulated or downregulated. Center: Venn diagrams illustrating the overlap of significantly up- or downregulated proteins identified in the two analyses. **(C)** Left: Associations between RS4DV and grey matter volume across 139 brain regions in disease-free participants, estimated using multiple linear regression (beta coefficients). Right: Genetic correlations between RS4DV and regional grey matter volumes (*r_g_*). Center: Scatter plot comparing region-wise associations at the phenotypic and genetic levels, with the Pearson correlation coefficient (*ρ*) and the whole-brain permutation-based *p*-value indicating the significance of their spatial correspondence. **(D)** Analogous analyses for white matter fractional anisotropy (FA) across major tracts. Statistical significance was evaluated using standard permutation testing rather than whole-brain spatial permutation. **(E)** Left: Serial mediation model (positive association protein model) linking PRS and brain structure in disease participants (*n* = 2,568). C’ = direct effect of PRS on brain structure; C = total effect of PRS on brain structure; total indirect effect = -0.004. Model fit indices: CFI = 0.95, RMSEA = 0.0. Right: Serial mediation model (negative association protein model) linking PRS and brain structure in disease participants. Total indirect effect = -0.001. Model fit indices: CFI = 0.94, RMSEA = 0.0. \**p* < 0.05, \*\*\**p* < 0.001.

We next examined molecular correlates of RS4DV using plasma proteomics in disease-free participants with proteomic data (*n* = 19,070) in the held-out test set. After control for the false discovery rate (FDR, *p_adj_* < 0.05), 1,063 proteins were positively and 222 negatively associated with RS4DV (Figure 4B). Moreover, 701 proteins differentiated individuals who later developed disease (*n* = 8,027) from those who remained zero diagnosis (*n* = 11,043), including 517 upregulated proteins and 184 downregulated proteins (Figure 4B). Upregulated proteins were enriched for inflammatory and metabolic markers, including IL-6, IL1RN, leptin, and FABP4, whereas downregulated proteins were involved in antioxidant defense and growth factor signaling (Supplementary Figure S16). These findings indicate that high RS4DV is characterized by systemic inflammation and metabolic dysregulation. Among the proteins found by the two analyses above, the most upregulated proteins (e.g., leptin, FABP4, IL1RN, ADM, HGF, FURIN, CPM and IL6) were linked to metabolic regulation^33–36^, inflammation^37,38^, tissue repair^39,40^ and immunity^38,41^, while the most downregulated proteins (e.g., PON3, CA14, IGFBP1/2, WFIKKN2) were implicated in oxidative stress defense^42^, acid-based regulation^43^, and growth factor signaling^44,45^. Enrichment analyses indicated that the overlap upregulated proteins of two sets were strongly involved in immune cell migration and cytokine activity, whereas the overlap downregulated proteins of two sets reflected pathways in lipid and hormone regulation (Supplementary Figure S15). The proteomic profile associated with high RS4DV was dominated by markers of systemic inflammation (IL-6, IL-1RN) and metabolic dysregulation (Leptin, FABP4).

To examine neural correlates, we linked the RS4DV to the brain in the disease-free individuals with structural MRI data in the held-out test set. RS4DV was significantly associated with grey matter (GM) volume across 67 regions (*n_GM_* = 18,286 with T1-weighted MRI data) and microstructural profile (fractional anisotropy, FA) in 40 white matter (WM, *n_WM_* = 17,402 with Diffusion MRI data) tracts (FDR, *p_adj_* < 0.05, Figure 4C-D). To assess biological consistency, we tested the genetic overlap between RS4DV and brain structure. Among the 67 structurally significant RS4DV-related brain regions, 13 showed significant genetic correlations (Supplementary Table S10). Spin tests confirmed that the overall spatial patterns of RS4DV-GM associations and genetic correlations were highly consistent (*ρ* = 0.500, *p*_spin_ = 0.005; Figure 4C). Although no single WM tract reached significant genetic correlation, permutation analyses validated that the spatial pattern of RS4DV-WM associations converged with genetic correlations across 48 tracts (*ρ* = 0.697, *p*_per_ < 0.001; Figure 4D).

Finally, we constructed a serial mediation model to map the flow of biological information among genotype, proteomics, RS4DV and brain phenotype. Among participants with baseline diagnosis, the positive association protein model revealed that the PRS had a significant indirect effect on GM volume via protein expression and RS4DV (*β* = - 0.004, 95% CI: [-0.006, -0.002], *p* < 0.001; Figure 4E, left). In contrast, no significant mediation effect was observed in the negative association protein model (Figure 4E, right). Among disease-free participants, no significant mediation effects were detected in either the positive or negative association protein models (Supplementary Figure S16).

### A light-version RS4DV model for real-world risk stratification

To facilitate practical implementation, we retrained a simplified RS4DV model based on six routinely available clinical and self-reported measures (Figure 5A), selected according to their importance in the full model and their accessibility in real-world settings. In the independent test cohort, the six-item model demonstrated preserved predictive performance (R² = 0.14, *p* < 0.001) and showed high consistency with the full model (*ρ* = 0.87, *p* < 0.001; Figure 5B), indicating that substantial dimensionality reduction did not largely compromise risk estimation.

**Figure 5.**
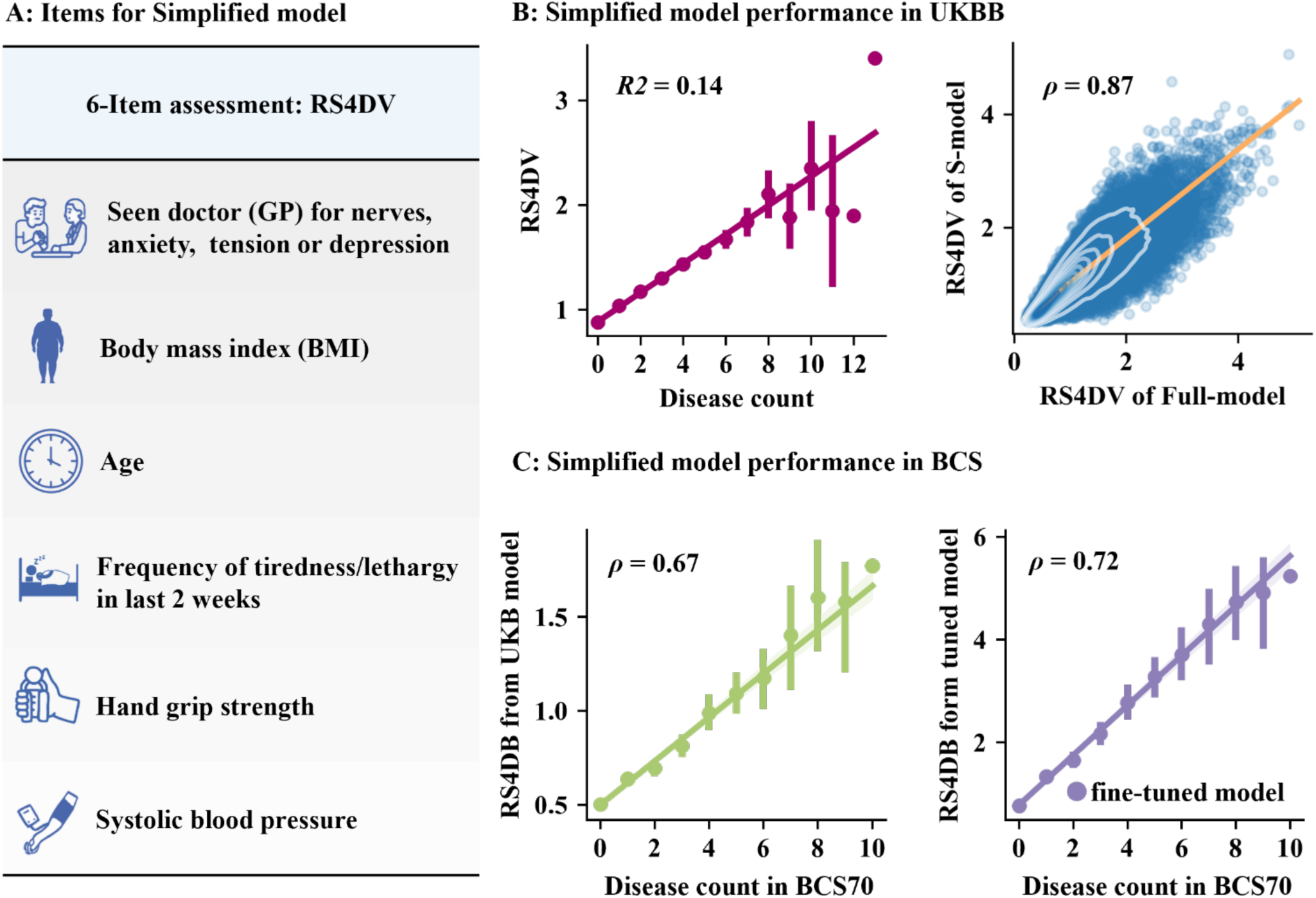
Performance of the simplified prediction model forRS4DV. **(A)** Schematic of the simplified model, which only used six key features of the full model to generate a concise 6-item assessment. **(B)** Left: Scatter plot showing the predictive performance of the simplified model in the test set (R² = 0.14, *p* < 0.001). Right: The RS4DV of simplified model (y-axis) was strongly correlated with those from the full model (x-axis), with a Pearson’s *ρ* = 0.87 (*p* < 0.001), indicating that the simplified model captured most of the variance explained by the full model. **(C)** Left: Predictive performance of the simplified model in an external cohort (BCS70), with a Pearson’s *ρ* = 0.65 (*p* < 0.001). Right: Performance of the fine-tuned simplified model in the same external cohort, achieving a Pearson’s *ρ* = 0.72 (*p* < 0.001), suggesting improved generalizability and robustness.

To further evaluate the generalizability of the simplified model, we validated it in the external 1970 British Birth Cohort (BCS70, *n* = 6,615), which was randomly divided into a training set (70%) and an independent test set (30%). We first applied the model trained in the UK Biobank directly to the BCS70 test set without additional optimization (zero-shot transfer, *n* = 1,973). Under this setting, the predicted scores showed a significant correlation with the observed outcomes (*ρ* = 0.67, *p* < 0.001), indicating substantial cross-cohort transferability. We further fine-tuned the pretrained model using the BCS70 training set. This adaptation led to improved model fit, resulting in higher predictive performance in the BCS70 test set (*ρ* = 0.72, *p* < 0.001; Figure 5C).

Consistent with the findings from the full model, Kaplan–Meier analyses based on the six-item RS4DV demonstrated that individuals in the top decile experienced significantly accelerated disease accumulation and higher mortality compared with those in the bottom decile (all log-rank *p* < 0.001). By year 15, in the validation subset, the cumulative incidence of disease progression reached 63.0% in the high-risk group versus 19.6% in the low-risk group, while the cumulative incidence of all-cause mortality was 10.4% versus 1.8%, respectively. In the test subset, the corresponding disease progression rates were 60.3% versus 22.3%, and mortality rates were 9.0% versus 1.0%. Similar and consistent patterns were observed when broader stratifications were applied (e.g., top 30% vs. bottom 30% of the distribution; Supplementary Figure S17). Furthermore, in the test subset, we compared individuals in the top 10% versus the bottom 10% of the 6-item RS4DV with analogous stratifications based on each of the six individual features. For disease progression outcomes, the 6-item RS4DV demonstrated superior risk discrimination compared with all six individual indicators. For all-cause mortality, the 6-item RS4DV showed a stratification performance comparable to age and outperformed the other five indicators (Supplementary Figure S18).

## Discussion

This study bridges a critical gap in preventive medicine by validating a scalable, BPS-informed metric for systemic multimorbidity risk. Grounded in the BPS framework, RS4DV frames disease vulnerability as a systemic health state shaped by the joint influence of biological, psychological, and social factors, rather than as the accumulation of isolated disease-specific risks. In contrast to conventional clinical risk scores that rely primarily on downstream pathological markers, RS4DV leverages upstream biopsychosocial determinants to identify latent vulnerability before the cascade of overt disease onset. Importantly, the score not only reflects current systemic health status but also prospectively predicts future disease progression among initially healthy individuals, enabling early stratification at a stage most amenable to primordial prevention. By integrating genetic, proteomic, and neuroimaging evidence, RS4DV captures shared biological pathways underlying multimorbidity, linking molecular mechanisms with convergent structural brain alterations. This convergence is consistent with a syndemics perspective, in which chronic diseases co-emerge through interacting biological pathways and shared sociopsychological drivers. This positions RS4DV as a potent tool for primordial prevention, enabling health systems to target interventions, such as social prescribing and lifestyle modification, at the root causes of health disparities^46^.

Previous studies have identified numerous factors associated with disease vulnerability or mortality risk, including disease-general factor^47,48^ and biological aging metrics derived from neuroimaging^49^ or plasma proteomics^50^. While these approaches have yielded valuable insights within individual domains, they largely remain siloed and do not explicitly model the cumulative and interactive nature of vulnerability emphasized by both the BPS model and the syndemics framework^8,9^. RS4DV addresses this limitation by integrating self-reported psychological traits (e.g., neuroticism, perceived stress), anthropometric measures (e.g., body mass index, grip strength), and lifestyle factors into a unified machine-learning framework that captures latent health deterioration across systems. Although RS4DV does not precisely predict the exact number of future diseases for each individual, it excels at ranking individuals according to their relative risk. This stratification capability provides a practical tool for identifying high-risk individuals and guiding early prevention and intervention, offering a multidimensional, interpretable, and scalable measure of health risk that complements prior domain-specific studies.

The reproducibility and clinical relevance of RS4DV were confirmed through rigorous validation. Analysis of the full model highlighted neuroticism, anthropometric measures, and life stress as the strongest contributors to cumulative disease vulnerability. Specifically, indicators such as prior consultation for nerves, anxiety, stress, or depression, along with key anthropometric measures including BMI and grip strength, emerged as early warning signals of disease progression. High BMI confers elevated risk for multiple chronic conditions, including diabetes, cardiovascular disease, and certain cancers^51^, whereas reduced grip strength reflects physical frailty and predicts disability, morbidity, and mortality^52^. Frequent tiredness or lethargy, capturing daily life stress, also contributed substantially to RS4DV prediction, consistent with evidence linking fatigue to adverse health outcomes^53^. Neuroticism further emerged as a key determinant, likely influencing disease vulnerability by shaping stress responses, health behaviours, and social adaptability^54^.

RS4DV enables robust and clinically relevant risk stratification for long-term health outcomes in initially healthy individuals. Although individual factors (e.g., neuroticism, life stress, BMI, and hand grip strength) are each associated with adverse outcomes, RS4DV shows substantially stronger risk separation than most single indicators in Kaplan–Meier analyses of disease progression and all-cause mortality over 14.7 years. This highlights that multimorbidity and premature death arise from the interplay of multiple BPS factors, underscoring the value of RS4DV as an integrated metric. Importantly, RS4DV incorporates both immutable factors (e.g., age) and modifiable factors (e.g., BMI, physical fitness, lifestyle behaviours, stress management), suggesting actionable targets for intervention. Recent works demonstrate that sparse plasma-protein signatures substantially enhance 10-year risk prediction across a broad spectrum of diseases^55^. Moreover, biological-age estimation via the proteomic age clock is strongly associated with multimorbidity and mortality, underscoring the growing consensus that preventive medicine urgently requires early, individualized, and quantitative indicators to forecast future disease vulnerability^56^. Our findings align closely with this paradigm, RS4DV serves as a precision metric that integrates mental, physical, and lifestyle dimensions into a unified risk framework. By identifying at-risk individuals before clinical onset, RS4DV bridges the gap between population-level risk factors and personalized preventive strategies, offering a tool for proactive health management and precision public health. High-risk individuals identified by RS4DV, while currently healthy, may benefit from lifestyle modifications such as increased physical activity, weight control, and stress reduction, thereby potentially mitigating future disease vulnerability.

Beyond its predictive utility, RS4DV sheds light on the molecular and neural mechanisms underlying multimorbidity. GWAS identified several genes, including MST1R, DNAJC27, and BANK1, which are not only involved in immune regulation and inflammatory signalling, but also associated with metabolic disorders such as obesity and diabetes^57,58^. Notably, both GWAS and proteomic enrichment analyses converged on the positive regulation of the MAPK cascade pathway, suggesting a coordinated role of this signalling axis in disease susceptibility. The RS4DV also showed significant genetic correlations with inflammation-related conditions such as obesity, diabetes, and depression, further supporting the involvement of inflammatory pathways. RS4DV was negatively associated with GM volume in the cerebellum and amygdala. The specific involvement of the amygdala, a hub for stress processing and emotional regulation is particularly notable^25,59^. Structural attrition in these regions suggests that systemic multimorbidity is paralleled by accelerated brain aging, potentially mediated by the same inflammatory cytokines identified in our proteomic analysis. This supports the existence of a bidirectional brain-body axis where central neural integrity and peripheral somatic health are mutually dependent^60^. Interestingly, the spatial association patterns of RS4DV’s with GM volume and WM microstructure are highly similar with the heritability maps of RS4DV with these brain features, suggesting a shared genetic basis. Overall, these findings demonstrate that RS4DV captures a measurable biological component of cumulative disease vulnerability, reflecting the interplay of genetic, molecular, and neural factors. Importantly, RS4DV also integrates environmental and lifestyle influences, highlighting its value as a multidimensional measure that links molecular and neural signatures with real-world health outcomes.

The development of light-version RS4DV aims to improve the real-world applicability of RS4DV in clinical and population settings. Although the full RS4DV provides a comprehensive assessment of disease vulnerability, its use may be limited in situations where time, data availability, or assessment burden are constrained. By reducing the model to six key items covering biological, psychological, and self-rated health domains, light-version RS4DV retains the core biopsychosocial structure while substantially improving feasibility. The selected items are routinely available clinical measures or simple self-reported indicators, allowing efficient implementation in primary care, large-scale screening, and remote health assessments. This simplified format supports integration into electronic health records, mobile health tools, and epidemiological surveys, where rapid risk stratification is often more practical than detailed phenotyping. From a preventive perspective, light-version RS4DV provides a practical tool for identifying individuals with elevated systemic vulnerability before the onset of overt multimorbidity. Its consistent performance in an independent cohort suggests robustness across populations. With further validation, light-version RS4DV may facilitate longitudinal monitoring and targeted intervention strategies in both public health and clinical practice.

Nevertheless, several limitations warrant consideration. First, we quantified baseline disease burden as an unweighted cumulative count of diagnoses recorded prior to recruitment. While this approach treats diverse conditions with equal weight and does not explicitly account for disease-specific severity or clinical staging, it serves as a pragmatic and robust proxy for an individual’s cumulative physiological toll. By capturing the aggregate burden, this metric provides a scalable summary of systemic health status across a large-scale population. In addition, the cohorts predominantly comprised individuals of European ancestry, and additional validation in ethnically and socioeconomically diverse populations is required to ensure generalizability. Although the proposed framework achieved good performance and stability, the current selection of representative diseases and BPS features may not fully capture the entire spectrum of disease vulnerability. Future expansion and refinement of this framework by incorporating broader disease categories, additional behavioural, environmental, and biological dimensions, and larger multi-ethnic cohorts, may further enhance its predictive utility and translational potential.

In conclusion, the RS4DV developed in this study provides a simple yet robust tool for predicting individual health trajectories, with meaningful ability to reflect differences across disease states, supported by convergent neural and molecular evidence. Notably, the light-version RS4DV, based on only six simple questions, enhances its feasibility for clinical and public health applications. Our findings highlight the potential of the RS4DV for early risk identification and its utility in supporting personalized health management and targeted interventions.

## Methods

### Study Population

This study utilized data from the UK Biobank^22^, a large-sample, ongoing prospective cohort that recruited over 500,000 participants in the United Kingdom (https://www.ukbiobank.ac.uk/). The information including demographics, lifestyle, physical characteristics, disease vulnerability at baseline or follow-up, biological samples and imaging data was recorded for the enrolled participants aged 40-70 years at recruitment. The baseline assessment (initial assessment visit, 2006-2010) and data collection via: (1) a touchscreen questionnaire, in which participants provided information on sociodemographic characteristics, lifestyle, psychosocial factors (e.g., social support and mental health), and medical history; (2) a face-to-face interview conducted by trained nurses to acquire early-life factors, employment status, medical conditions, medications, and surgical history; (3) physical measurements; and (4) biological sampling (blood assays, saliva assays and urine assays). In this study, the data of 502,180 participants were available at the time of our request. The UK Biobank data obtained ethical approval from the North West Multi-Center Research Ethics Committee (reference number 16/NW/0274), and all participants provided informed consent electronically.

### Risk score for disease vulnerability (RS4DV) modelling

#### Diseases cumulation

To construct the RS4DV model, we defined cumulative disease as the simple count of diagnosed conditions from a predefined list. This approach provides a straightforward, overall measure of an individual’s health status, without assuming differential clinical severity across conditions. This approach was chosen to create a general index of an individual’s overall health status rather than to capture comorbidity. We quantified the disease vulnerability for each participant using a diseases list that included physical conditions (e.g., stroke, asthma, dermatitis, arthritis, dorsalgia, ulcer, colitis, irritable bowel syndrome, hearing loss, visual disturbances, obesity, insulin-dependent and non-insulin-dependent diabetes mellitus, essential and secondary hypertension, hernia, bronchitis, movement disorders, kidney and ureteral calculi) as well as mental disorders (e.g., conduct disorder, depression, other anxiety disorders, phobia, pervasive developmental disorders, obsessive-compulsive disorder, schizophrenia, bipolar disorder, psychosis, manic episode, and adjustment disorders). Diseases were defined and classified based on International Classification of Diseases (ICD-10) codes. The participants were classified as having a disease if the first reported diagnosis occurred before the baseline assessment. The field of first occurrences (Category 1712) in the UK Biobank Health-related Outcomes dataset was used to extract the disease status of each subject. The incidence rates for each disease based on ICD-10 codes in the UK Biobank data are provided in Supplementary Table S1.

#### BPS features selection

A total of 85 BPS features collected by touchscreen assessment at the baseline were selected for modelling individual RS4DV, as these features were previously demonstrated to capture key determinants of chronic health outcomes^61^ and were harmonized by excluding redundant and poorly measured variables. These BPS features are categorized into three domains: mental health, physical health, and sociodemographic factors. The mental health included three subcategories: (1) Neuroticism (individual items and total score) based on 12 behaviours closely associated with negative affect; (2) Trauma (occurrences of disease, injury, bereavement, or stress in the past 2 years), comprising six events; (3) Emotional factors, including the frequency of certain emotions reported in the past 2 weeks and consultations with general or psychiatric practitioners for issues related to neuroticism, anxiety, tension, or depression. The physical health comprised four subcategories: (1) Physical activity assessed using the MET score calculated from the International Physical Activity Questionnaire (IPAQ); (2) Sleep (questions regarding sleep duration, daytime napping, snoring, and insomnia); (3) Substance use (e.g., smoking and alcohol consumption); (4) Anthropometric measurements (e.g., body mass index (BMI), fractures over the past 5 years, and blood pressure). The sociodemographic factors included three subcategories: (1) Socioeconomic status (e.g., educational attainment, income, and employment status); (2) Occupational indicators (e.g., household composition, social companionship, and engagement in manual labor); (3) Demographics (e.g., age, sex, and ethnicity). A complete list of these BPS factors and their corresponding UK Biobank data fields is provided in Supplementary Table S3.

#### Data preprocessing

To ensure data quality, participants with more than 20% missing values across the 85 BPS features were excluded from the full UK Biobank baseline sample (*n* = 502,180), leaving 482,057 individuals for analysis. These individuals were split into a discovery cohort (*n* = 391,193), which did not include follow-up proteomic or imaging data, and a test cohort (*n* = 90,864) that contained these modalities for downstream analyses. In line with previous research, Remaining item-level missingness was addressed in the primary analysis using a data-driven Bayesian ridge regression imputation in Scikit-learn^62^, each missing value was modelled as a function of the other features, with the feature-wise median used to initialize the imputation^61^. Imputation was performed separately within the discovery and test cohorts to avoid information leakage. To evaluate the reliability of this imputation, we performed a sensitivity analysis comparing model predictions obtained from models trained on the Bayesian-imputed data with predictions from models trained on complete-case analysis (CCA)^63–67^; agreement between predicted scores was quantified using Pearson correlation.

#### RS4DV Modelling

To develop a personalized RS4DV, we employed 85 BPS features to predict individuals’ cumulative disease count. The discovery cohort (*n* = 391,193) was randomly partitioned into a training set (*n* = 273,835) and a validation set (*n* = 117,358) in a 7:3 ratio, while an independent test cohort (*n* = 90,864) was reserved for external evaluation. Initially, we compared the predictive accuracy of five commonly used algorithms, including Light Gradient Boosting Machine (LGBM)^68^, Support Vector Regression (SVR)^69,70^, Random Forest (RF)^71^, Partial Least Squares (PLS)^72^, and Ridge Regression^73^, using their default hyperparameter settings on the independent test cohort. Consistent with algorithmic requirements, continuous variables were standardized for SVR, Ridge Regression, and PLS, while tree-based models (LGBM and RF) utilized non-standardized data. Model performance was evaluated using the coefficient of determination (R^2^), mean squared error (MSE), and mean absolute error (MAE). After identifying the algorithm with the superior baseline performance, we used LGBM to construct the final RS4DV model. Specifically, grid search combined with five-fold cross-validation was performed within the training set to optimize hyperparameters. During model training, the validation set was incorporated as an independent evaluation dataset for early stopping: model performance on the validation set was monitored at each boosting iteration, and training was terminated when no improvement was observed for a predefined number of consecutive iterations, thereby preventing overfitting.

#### Model evaluation

We first evaluated the overall performance of the optimized model in the independent test cohort (*n* = 90,864, with proteomic and imaging data) using R^2^ and root mean squared error (RMSE). Feature importance was assessed using tree-based methods, including information gain and split frequency. For model interpretation, SHapley Additive exPlanations (SHAP) were used to quantify feature contributions at both global and individual levels. Global feature importance was defined by integrating multiple complementary importance metrics derived from the final LGBM model, including (i) gain-based importance (reflecting the total loss reduction attributed to a feature), (ii) split-based importance (capturing the frequency with which a feature was used for tree splitting), and (iii) SHAP-based importance (defined as the mean absolute SHAP value across all individuals, representing the average marginal contribution of each feature to the model output). To ensure comparability across metrics, all three importance measures were independently normalized to a [0,1] range using min–max scaling. A composite importance score was then calculated as a weighted sum of the normalized metrics, with weights of 0.25 for gain importance, 0.25 for split importance, and 0.5 for SHAP importance. This weighting scheme prioritized SHAP values due to their solid theoretical foundation in cooperative game theory and their ability to provide consistent, model-agnostic estimates of feature contributions^74,75^. Features were subsequently ranked according to this combined importance score to identify the most influential predictors driving RS4DV. To further investigate the relative contributions of different feature domains, the 85 features were grouped into three domains (physical health, mental health, and sociodemographic factors). Models were trained using features from one domain, any two domains, or all three domains, and their R^2^ values in the test set were compared. In addition, features were categorized into 10 subdomains, and the significance of each subdomain was assessed by comparing its observed R² with a null distribution generated from 10,000 permutations; subdomains with *p* < 0.05 were considered significant.

### Survival analysis

#### Kaplan-Meier curves

To evaluate whether the RS4DV could predict disease progression and all-cause mortality, individuals were divided into high- and low-risk groups based on the top and bottom 10% of the RS4DV distribution. Kaplan-Meier (KM) curves^76,77^ were constructed to visualize survival differences between high- and low-risk groups in both disease progression and all-cause mortality, and statistical significance was assessed using the log-rank test, with a significance threshold at *p* < 0.05. The median follow-up time estimated using the reverse Kaplan-Meier method. All survival analyses were conducted in Python using the lifelines package^78^. For validation, an alternative grouping using the top and bottom 30% of the distribution was also applied.

#### Cox proportional hazards regression

To further assess the associations between RS4DV and the occurrences of disease progression and all-cause mortality, we performed univariate Cox proportional hazards regression analyses^79^. RS4DV was entered into Cox model to evaluate its respective predictive effects on disease progression and all-cause mortality. Time-to-event was defined as the time from baseline to the first occurrence of disease or the end of follow-up (September 2023), whichever occurred first. Hazard ratios (HRs) and corresponding 95% confidence intervals (CIs) were reported, and statistical significance was set at *p* < 0.05. All survival analyses were conducted in Python using the lifelines package^78^.

### Follow-up Disease Incidence Analysis

To evaluate the stratification ability of RS4DV for individual diseases during the follow-up period, participants were classified into high-risk and low-risk groups based on the top 10% and bottom 10% of the RS4DV distribution, respectively. For each group, we calculated the frequency of incident cases for 30 pre-specified diseases. Disease was defined during the follow-up among participants without the disease at baseline. Frequencies were summarized using counts and percentages, and comparisons between high- and low-risk groups. All analyses were conducted separately in the validation and test subsets to assess the robustness and reproducibility of RS4DV’s predictive stratification.

### Proteomic analyses

#### Protein data

To identify the RS4DV associated proteins, multivariable regression analysis and logistic regression analysis were conducted. In the UK Biobank Pharma Proteomics Project (UKB-PPP) consortium, plasma samples collected from 54,219 UK Biobank participants were analysed using the Olink Explore™ Proximity Extension Assay and next-generation sequencing^80^. This analysis measured 2,941 protein analytes corresponding to 2,923 unique proteins (see details in biobank.ndph.ox.ac.uk/ukb/ukb/docs/PPP_Phase_1_QC_dataset_companion_doc.pdf). After excluding proteins with more than 10% missing data, a total of 1,459 proteins were included in our study.

#### Multivariable regression analysis

To investigate the molecular basis of RS4DV, we performed separate multivariable linear regression analyses to examine the associations between RS4DV and each of the 1,459 protein phenotypes, with sex and age included as covariates in each model. The primary analysis used the Benjamini-Hochberg false discovery rate (FDR-BH) procedure with a significance threshold of 0.05. For sensitivity analysis, a more stringent Bonferroni correction was applied, with a significance level set at *p* < 0.001.

#### Logistic regression analysis

To further examine whether the proteins significantly associated with RS4DV also exhibited differential expression between individuals who later develop the disease (future converters) and those who remain healthy throughout the study (stable healthy controls, defined as individuals without the disease at both baseline and follow-up), we conducted logistic regression analyses based on the proteins identified in the above multivariable linear regression analyses. The logistic regression analysis was adjusted for sex and age, and the primary analysis used the FDR-BH procedure with a significance threshold of 0.05. For sensitivity analysis, a more stringent Bonferroni correction was applied, with a significance level set at *p* < 0.001.

### Neuroimaging analyses

#### MRI data

We utilized multimodal brain imaging data, encompassing both brain T1 structural MR images and diffusion MR images, to identify the associations between the RS4DV and the regional GMV and microstructure of WM tracts. Data acquisition was conducted using a standard Siemens Skyra 3T scanner with a standard 32-channel RF receiver head coil. The pre-processed region-wise GMV and tract-wise FA data were provided by UK Biobank. Specifically, to assess regional GMV, UK Biobank employed the FAST algorithm in the FSL toolkit (https://fsl.fmrib.ox.ac.uk/fsl). Using FAST, the GMV map was generated for each participant. Then, the whole brain was segmented into 139 regions of interest (ROIs) using the Harvard-Oxford cortical and subcortical atlases and the Diedrichsen cerebellar atlas. The GMV of each ROI was obtained from Category 1101 in UK Biobank. For WM tracts, we selected mean fractional anisotropy (FA) values to characterize WM microstructure. To avoid partial volume effects and registration errors, the whole-brain FA map of each subject was first aligned onto a standardized WM skeleton. Then, the whole-brain WM was segmented into 48 tracts using the ICBM-DTI-81 white matter atlas and overlaid the FA map on the skeleton^81–83^. The mean FA value of each WM tract on the skeleton was calculated for each participant. The mean FA value of each WM was obtained from Category 134 in UK Biobank.

#### Multivariable regression analysis

To explore the neural basis underlying RS4DV, we conducted separate multivariable linear regression analyses to assess the associations between RS4DV and GMV of 139 GM regions and FA of 48 WM tracts, including sex and age as covariates in each model. Additionally, total GM volume was included as a covariate when analysing regional GM volumes, and scanner site was considered as a covariate for all neuroimaging analyses. An FDR-BH correction was applied for multiple comparisons, with a significance threshold of *p* < 0.05.

### Genomic analysis

#### Genotypes, quality control, and principal component analysis (PCA)

The majority of the UK Biobank samples were genotyped using the Affymetrix UK Biobank Axiom array (Santa Clara, CA, USA) with the rest of the samples genotyped using the Affymetrix UK BiLEVE Axiom array. The details for quality control, phasing and imputation are described in the following linkages (http://biobank.ctsu.ox.ac.uk/crystal/refer.cgi?id=155583; http://biobank.ctsu.ox.ac.uk/crystal/refer.cgi?id=155580; http://biobank.ctsu.ox.ac.uk/crystal/refer.cgi?id=157020). In this study, individuals were included with genetically inferred to be of White British ancestry, without sex chromosome aneuploidy, and unrelated within the third degree. Only participants with autosomal genotype data and those contributing to the calculation of genetic principal components were retained. At the variant level, single nucleotide polymorphisms (SNPs) meeting the following quality control criteria were kept: minor allele frequency (MAF) > 0.005, Hardy-Weinberg equilibrium (HWE) *p* > 1×10⁻⁶, and genotype missingness < 5%. To ensure genotype accuracy and cross-dataset compatibility, only SNPs from the HapMap 3 reference panel were included. After quality control, 1,153,470 SNPs and 336,950 participants remained. PCA was performed to capture population structure. For subsequent analyses, we only used the subset of healthy individuals (*n* = 126,950) who had no recorded diseases prior to baseline assessment.

#### GWAS and linkage disequilibrium score regression (LDSC)

To investigate the genetic associations for the RS4DV, we conducted a GWAS using the Genome-wide Complex Trait Analysis (GCTA) tool^84^ in the healthy individuals. The Haseman-Elston regression method was used to estimate heritability^85^. Then, a mixed-model analysis was performed using fastGWA^86^ with age, sex, and the top 4 genetic principal components as covariates, and 1,000 null SNPs for parameter adjustment. To further investigate the genetic architecture of RS4DV and assess potential confounding, we conducted LDSC analysis. Because the genomic inflation factor (λ_GC_) increases with sample size even in the absence of confounding^87^, the LDSC was used to distinguish true polygenic signal from confounding bias to improve the interpretability of GWAS results.

#### Polygenic risk score (PRS) for RS4DV

To evaluate the heritable component of RS4DV and provide a quantitative measure, we derived the PRS as an aggregate measure of genetic susceptibility. The SNPs used for PRS calculation were selected from the GWAS results of RS4DV. To account for linkage disequilibrium (LD), we applied LD-based clumping using PLINK 2.0, and the SNPs with at least 250 kilobases apart, pairwise LD (*r*²) < 0.2, and a significance threshold of *p* < 5×10⁻⁵ were retained for analysis. PRS for each individual was calculated as the weighted sum of risk allele dosages, with weights corresponding to GWAS-derived effect sizes. Finally, the resultant PRS was standardized using z-transformation to reduce inter-individual variability.

We also computed PRS for patients who had reported at least one disease prior to baseline assessment. The patients were classified into three subtypes based on disease progression after baseline: no progression, mild progression (1 ≤ number of diseases ≤ 2), and severe progression (number of diseases ≥ 3). One-way analysis of variance was performed to determine the difference in PRS among the three subtypes with significant level set at *p* < 0.05. Post-hoc two-sample t-tests were conducted to identify the between groups difference, with statistical significance set at *p* < 0.05 corrected with Bonferroni method.

#### Genetic correlations analysis

To evaluate the genetic correlations between RS4DV and a wide range of phenotypes, we applied bivariate LD score regression (LDSC)^88^. Summary statistics for disease phenotypes were obtained from the Finngen R12 GWAS results^89^. For 46 behavioural and health-related traits as well as 187 brain imaging phenotypes (including grey matter volume of 139 regions and fractional anisotropy of 48 white matter tracts), we used GWAS results reported by Jiang et al.^86,90^, derived from genome-wide genotyping data of 456,422 individuals of European ancestry in the UK Biobank. LDSC leverages the correlation structure of SNPs due to LD to estimate the shared genetic architecture between two traits, and is robust to sample overlap. Multiple comparisons were corrected using the FDR-BH, with a significance threshold of *p* < 0.05.

To investigate the correlations between the beta maps directly associated with RS4DV of the brain imaging measures and the genetic correlation maps of RS4DV with brain phenotypes, we assessed their spatial correlations to eliminate spatial autocorrelation through permutation tests. When assessing spatial correlations between brain maps, the inherent spatial smoothness of surfaces can inflate the apparent statistical significance. To address this issue, we employed a spin permutation test to evaluate the statistical significance of whole brain maps using BrainSMASH^91^. Specifically, the spatial coordinates of each brain region were projected onto a spherical surface to approximately preserve their topological arrangement. The surface was then randomly rotated multiple times, and the original brain map values were reassigned to the rotated coordinates. This process generated a null distribution that maintained the spatial autocorrelation structure. The observed Pearson correlation coefficient between brain maps was subsequently compared to the empirical distribution to determine significance. For white matter tracts, whose geometry does not permit spherical projection and rotation, we used a classical non-parametric permutation test. This involved randomly shuffling the correspondence between the two data vectors to generate a null distribution of correlation coefficients. The empirical *p*-value was then calculated by comparing the observed statistic to this distribution.

### Serial mediation model analyses

Given the temporal structure of our data, in which genetic/PRS data preceded baseline assessments (protein and RS4DV) and these, in turn, preceded later-life brain imaging, we hypothesized a potential causal cascade. To examine this, we constructed a chain mediation model to test whether the PRS influences brain structure through alterations in RS4DV mediated by protein expression. Specifically, the total volume of GM regions that showed significant associations in both multivariable linear regression and genetic association analyses was calculated as an indicator of GM alteration. For protein expression, we separately aggregated the levels of significantly upregulated and downregulated proteins identified in proteomic analyses to represent overall protein-level changes.

Within the structural equation modelling (SEM) framework, we specified paths from PRS to protein expression, from protein expression to RS4DV, and from both protein expression and RS4DV to GM volume. A direct path from PRS to GM volume was also included. The model was estimated using maximum likelihood, and bootstrap resampling with 5000 iterations was performed to obtain robust standard errors and confidence intervals for the indirect effects. To enhance the robustness and generalizability of the findings, the same analyses were repeated in two subgroups: participants who were disease-free at recruitment and a separate patient cohort. This allowed us to assess the consistency of the mediation pathways across healthy and clinical populations. Statistical significance was defined as *p* < 0.05. All analyses were conducted using the lavaan^92^ package in R.

### Simplified model for RS4DV

#### Replication cohorts overview

To test the generalization of RS4DV, we incorporated a new external dataset, BCS70, for the generalization test. The dataset of BCS70 is a major longitudinal cohort in the United Kingdom that originally collected data of all infants born in 1970^93^, with multiple follow-up assessments to track changes in health, education, employment, and socioeconomic status throughout the life course. In the present study, we specifically used data from the age 46 sweep. The dataset of BCS70 refers to the sixth follow-up of the 1970 British Cohort Study (conducted at age 46). After excluding participants with missing data (including responses marked as "don’t know" or "no answer"), 8,085 participants in BCS70 dataset were used for analysis. In the BCS70 follow-up data, some new information was added: (1) Disease vulnerability (including chronic diseases, lifestyle factors, and anthropometric measurements); (2) Mental health (e.g., scales for anxiety, depression, and life satisfaction); (3) Cognitive function (e.g., cognitive and memory tests); (4) Lifestyle and environmental factors (e.g., diet, physical activity, smoking, and alcohol consumption); and (5) Socioeconomic status (e.g., educational attainment, employment, and income) as well as family and social relationships.

#### Replication disease phenotypes

In the BCS70 dataset, we selected a subset of diseases from the UK Biobank diseases list due to differences in disease coverage between the two cohorts, including 14 physical diseases (obesity, diabetes, stroke, visual impairments, hearing impairments, hypertension, asthma, ulcer, irritable bowel syndrome, eczema, arthritis, kidney/bladder stones, and back prolapsed disk sciatica) and 5 mental illnesses (anxiety, major depressive disorder, phobia, schizophrenia, and obsessive-compulsive disorder). Participants were asked whether they had ever been diagnosed with a given condition, and an affirmative response was considered indicator of disease presence.

#### Light-RS4DV modelling

To enhance the practical applicability of RS4DV, we developed a simplified model (Light-RS4DV) by reducing the number of input features. Feature selection was conducted based on three importance metrics derived from the optimal machine-learning model: information gain ranking, feature split frequency, and average SHAP value ranking. The six features with the highest aggregated importance scores among the original 85 features were selected and used to re-train the RS4DV model. The Light-RS4DV model was trained using these selected features in the original training set and subsequently evaluated in the test set to assess its performance. To further examine generalizability, the model was tested in an independent external cohort (BCS70 dataset). In addition, a transfer learning–based fine-tuning strategy was applied to improve model adaptability in the external cohort. Specifically, the pretrained Light-RS4DV model trained on the original dataset was used to initialize the model parameters, after which the model was further trained on the BCS cohort using the same feature set, allowing the model to adjust its learned representations to the target population while retaining information from the source dataset.

## Data availability

This research was conducted using data from the UK Biobank under access application number 197910. UK Biobank makes its data available to all bona fide researchers for any health-related research that serves the public interest, without granting preferential or exclusive access to any individual or group. All researchers are subject to the same application process and approval criteria, as defined by UK Biobank. For further details regarding the access procedure, please visit the UK Biobank website: http://www.ukbiobank.ac.uk/register-apply/.

## Code availability

Simplified model for RS4DV have been deposited at https://huggingface.co/spaces/Jtmcute/RS4DV. Code is available at https://github.com/jiaojianwang/RS4DV.

## Acknowledgement

This work was supported by the National Natural Science Foundation of China (82425024) and the Yunnan Fundamental Research Projects (202501AV070005, 202201BE070001-004). Additional support was provided to Prof. Samuele Cortese, NIHR Research Professor (NIHR303122), by the National Institute for Health and Care Research (NIHR). The views expressed in this publication are those of the author(s) and not necessarily those of the NIHR, NHS, or the UK Department of Health and Social Care. Prof. Cortese is also supported by NIHR grants NIHR203684, NIHR203035, NIHR130077, NIHR128472, RP-PG-0618-20003, and by grant 101095568-HORIZONHLTH-2022-DISEASE-07-03 from the European Research Executive Agency.

## Author contributions

J. Wang, B. Liu and C. Chu contributed to the conception and design of the study. J. Chen contributed to data processing and analysing. J. Chen, B. Liu, C. Chu, and J. Wang wrote and edited the manuscript. C. Chu, B. Liu, J. Wang, S. Cortese, M. Garcia-Argibay, C. Christogiannis and W. Li provided significant feedback on the analyses and the manuscript. J. Chen produced the figures. All the authors commented and discussed the results.

## Compliance with Ethical Standards

This study used a public dataset which obtained the ethical improvement.

