## Supplementary Materials: Figures and Tables for "A Biopsychosocial Risk Score for Stratifying Disease Vulnerability in Healthy Populations: A Prospective Cohort and Multi-Omics Study in the UK Biobank"

**Catalogues**

#### Supplementary Tables

**Table S1. Health conditions**

| Disorders | ICD-10 | Number of cases | Chapter | Major disease categories | Field ID |
|---|---|---|---|---|---|
| Conduct disorder | F91 | 103 | Chapter V | Mental and behavioural disorders | 130978 |
| Depression | F32 | 41,246 | Chapter V | Mental and behavioural disorders | 130894 |
| Other anxiety | F41 | 18,138 | Chapter V | Mental and behavioural disorders | 130906 |
| Phobia anxiety | F40 | 1,568 | Chapter V | Mental and behavioural disorders | 130904 |
| Pervasive developmental disorders | F84 | 44 | Chapter V | Mental and behavioural disorders | 130970 |
| OCD | F42 | 625 | Chapter V | Mental and behavioural disorders | 130908 |
| Schizophrenia | F20 | 912 | Chapter V | Mental and behavioural disorders | 130874 |
| Bipolar | F31 | 1,736 | Chapter V | Mental and behavioural disorders | 130892 |
| Psychosis | F29 | 223 | Chapter V | Mental and behavioural disorders | 130888 |

| Disorders | ICD-10 | Number of cases | Chapter | Major disease categories | Field ID |
|---|---|---|---|---|---|
| Manic episode | F30 | 319 | Chapter V | Mental and behavioural disorders | 130890 |
| Adjustment disorders | F43 | 10,370 | Chapter V | Mental and behavioural disorders | 130910 |
| Stroke | I64 | 7,032 | Chapter IX | Circulatory system disorders | 131368 |
| Asthma | J45 | 59,172 | Chapter X | Respiratory system disorders | 131494 |
| Dermatitis | L20, L21, L22, L23, L24, L25, L26, L27, L30 | 47,974 | Chapter XII | Skin and subcutaneous tissue disorders | 131720, 131722, 131724, 131726, 131728, 131730, 131732, 131734, 131740 |
| Arthritis | M00, M05, M06, M08, M13 | 19,404 | Chapter XIII | Musculoskeletal system and connective tissue disorders | 131840, 131848, 131850, 131854, 131864 |
| Dorsalgia | M54 | 47,721 | Chapter XIII | Musculoskeletal system and connective tissue disorders | 131928 |
| Ulcer | K25, K26, K27, K28 | 12,784 | Chapter XI | Digestive system disorders | 131590, 131592, 131594, 131596 |
| Colitis | K51, K52 | 16,880 | Chapter XI | Digestive system disorders | 131628, 131630 |
| IBS | K58 | 25,794 | Chapter XI | Digestive system disorders | 131638 |

| Disorders | ICD-10 | Number of cases | Chapter | Major disease categories | Field ID |
|---|---|---|---|---|---|
| Hearing loss | H90, H91 | 8,862 | Chapter VIII | Ear and mastoid process disorders | 131258, 131260 |
| Visual disturbances | H53, H54 | 6,109 | Chapter VII | Eye and adnexa disorders | 131210, 131212 |
| Obesity | E66 | 13,524 | Chapter IV | Endocrine, nutritional and metabolic diseases | 130792 |
| Insulin-dependent diabetes mellitus | E10 | 2,648 | Chapter IV | Endocrine, nutritional and metabolic diseases | 130706 |
| Non-insulin-dependent diabetes mellitus | E12 | 13,600 | Chapter IV | Endocrine, nutritional and metabolic diseases | 130708 |
| Essential (primary) hypertension | I10 | 134,417 | Chapter IX | Circulatory system disorders | 131286 |
| Secondary hypertension | I15 | 141 | Chapter IX | Circulatory system disorders | 131294 |
| Hernia | K40, K41, K42, K43, K44, K45, K46 | 45415 | Chapter XI | Digestive system disorders | 131612, 131614, 131616, 131618,131620,131622,131624,131634 |

| Disorders | ICD-10 | Number of cases | Chapter | Major disease categories | Field ID |
|---|---|---|---|---|---|
| Bronchitis | J20, J40, J41, J42 | 11493 | Chapter X | Respiratory system disorders | 131458, 131484, 131486, 131488 |
| Movement disorders | G25 | 1884 | Chapter VI | Nervous system disorders | 131032 |
| Calculus of kidney and ureter | N20 | 7049 | Chapter XIV | Genitourinary system disorders | 7049 |

**Table S2. UKBB participants used in the specific statistical analyses**

| Dataset | Instance | Sample size |
|---|---|---|
| Touchscreen | Instance 0 | Drop more than 20% of features missing in **85** features (**482,057**)<br>Trian and Test set (**391,193**)<br>Validation set (**90,864**) |
| Protein data | Instance 0 | **50,819** |
| T1 structural brain MRI | Instance 2 | **45,963** |
| Diffusion brain MRI | Instance 2 | **43,677** |

**Table S3. RS4DV indicators**

| Field id | Feature | Type | Cluster |
|---|---|---|---|
| 31 | Sex | Sociodemographic | Demographic |
| 102 | Pulse rate | Physical health | Anthropometric |
| 709 | Number in household | Sociodemographic | Socioeconomic |
| 728 | Number of vehicles in household | Sociodemographic | Socioeconomic |
| 738 | Average total household income | Sociodemographic | Socioeconomic |
| 806 | Job involves walking or standing | Sociodemographic | Occupational |
| 816 | Job involves heavy manual or physical work | Sociodemographic | Occupational |
| 1031 | Frequency of friends and family visits | Sociodemographic | Socioeconomic |
| 1160 | Sleep duration | Physical health | Sleep |
| 1170 | Getting up in the morning | Physical health | Sleep |
| 1180 | Chronotype | Physical health | Sleep |
| 1190 | Nap during the day | Physical health | Sleep |
| 1200 | Sleeplessness/insomnia | Physical health | Sleep |
| 1220 | Daytime dozing (narcolepsy) | Physical health | Sleep |
| 1249 | Past tobacco smoking | Physical health | Substance use |
| 1259 | Smokers in household | Physical health | Substance use |
| 1269 | Hours of exposure to tobacco at home | Physical health | Substance use |
| 1558 | Alcohol intake frequency | Physical health | Substance use |
| 1628 | Alcohol intake vs. 10 years previously | Physical health | Substance use |
| 1920 | Mood swings | Mental health | Mood |
| 1930 | Miserableness | Mental health | Mood |
| 1940 | Irritability | Mental health | Mood |
| 1950 | Sensitivity/hurt feelings | Mental health | Mood |
| 1960 | Fed-up feelings | Mental health | Mood |
| 1970 | Nervous feelings | Mental health | Mood |
| 1980 | Worrier/anxious feelings | Mental health | Mood |
| 1990 | Tens/highly strung | Mental health | Mood |
| 2000 | Worry too long after embarrassment | Mental health | Mood |
| 2010 | Suffer from nerves | Mental health | Mood |
| 2020 | Loneliness | Mental health | Mood |
| 2030 | Guilty feeling | Mental health | Mood |
| 2040 | Risk taking | Mental health | Life stressors |
| 2050 | Frequency of depressed mood in last 2 weeks | Mental health | Life stressors |

| Field id | Feature | Type | Cluster |
|---|---|---|---|
| 2060 | Frequency of unenthusiasm/disinterest in last 2 weeks | Mental health | Life stressors |
| 2070 | Frequency of tenseness/restlessness in last 2 weeks | Mental health | Life stressors |
| 2080 | Frequency of tiredness/lethargy in last 2 weeks | Mental health | Life stressors |
| 2090 | Seen doctor (GP) for nerves, anxiety, tension or depression | Mental health | Neuroticism |
| 2100 | Seen psychiatrist for nerves, anxiety, tension or depression | Mental health | Neuroticism |
| 2110 | Able to confide | Sociodemographic | Occupational |
| 2306 | Weight change compared with one year ago | Physical health | Anthropometric |
| 2463 | Fractured/broken bones in last 5 years | Physical health | Anthropometric |
| 4079 | Diastolic blood pressure | Physical health | Anthropometric |
| 4080 | Systolic blood pressure | Physical health | Anthropometric |
| 6145 | Serious illness, injury, bereavement in the last 2 years | Mental health | Life stressors |
| 6160 | Leisure/social activities | Sociodemographic | Socioeconomic |
| 20116 | Smoking status | Physical health | Substance use |
| 20117 | Alcohol drinker status | Physical health | Substance use |
| 20160 | Ever smoked | Physical health | Substance use |
| 21001 | Body mass index (BMI) | Physical health | Anthropometric |
| 21002 | Weight | Physical health | Anthropometric |
| 21003 | Age | Sociodemographic | Demographic |
| 22032 | IPAQ activity group | Physical health | Physical activity |
| 22035 | Above moderate/vigorous recommendation | Physical health | Physical activity |
| 22037 | MET minutes per week for walking | Physical health | Physical activity |
| 22038 | MET minutes per week for moderate activity | Physical health | Physical activity |
| 22039 | MET minutes per week for vigorous activity | Physical health | Physical activity |
| 46&47 | Hand grip strength | Physical health | Anthropometric |
| 21000 | White | Sociodemographic | Demographic |
| 21000 | Mixed | Sociodemographic | Demographic |
| 21000 | Asian or Asian British | Sociodemographic | Demographic |

| Field id | Feature | Type | Cluster |
|---|---|---|---|
| 21000 | Black or Black British | Sociodemographic | Demographic |
| 21000 | Chinese | Sociodemographic | Demographic |
| 21000 | Other ethnic group | Sociodemographic | Demographic |
| 6138 | CSEs or equivalent | Sociodemographic | Demographic |
| 6138 | College or University degree | Sociodemographic | Demographic |
| 6138 | NVQ or HND or HNC or equivalent | Sociodemographic | Occupational |
| 6138 | A levels/AS levels or equivalent | Sociodemographic | Occupational |
| 6138 | O levels/GCSEs or equivalent | Sociodemographic | Occupational |
| 6138 | Other professional qualifications eg: nursing, teaching | Sociodemographic | Occupational |
| 6138 | Total number of qualifications reported | Sociodemographic | Occupational |
| 6142 | Unable to work because of sickness or disability | Sociodemographic | Occupational |
| 6142 | In paid employment or self-employed | Sociodemographic | Occupational |
| 6142 | Unemployed | Sociodemographic | Occupational |
| 6142 | Retired | Sociodemographic | Occupational |
| 6142 | Looking after home and/or family | Sociodemographic | Occupational |
| 6142 | Doing unpaid or voluntary work | Sociodemographic | Occupational |
| 6142 | Full or part-time student | Sociodemographic | Occupational |
| 6141 | Live with other related | Sociodemographic | Socioeconomic |
| 6141 | Live with grandchild | Sociodemographic | Socioeconomic |
| 6141 | Live with other unrelated | Sociodemographic | Socioeconomic |
| 6141 | Live with grandparent | Sociodemographic | Socioeconomic |
| 6141 | Live with brother and/or sister | Sociodemographic | Socioeconomic |
| 6141 | Live with son and/or daughter (include step-children) | Sociodemographic | Socioeconomic |
| 6141 | Live with mother and/or father | Sociodemographic | Socioeconomic |
| 6141 | Live with Husband, wife or partner | Sociodemographic | Socioeconomic |

**Table S4. Baseline Characteristics**

| Characteristic | Discovery Cohort (n=391,193) | Test Cohort (n=90,864) | SMD (Standardized Mean Difference) |
|---|---|---|---|
| Age, mean (SD) | 56.53 (8.11) | 55.98 (7.95) | 0.0675 |
| Female, n (%) | 213,108 (54.4%) | 48,440 (53.6%) | 0.0234 |
| **Multiple Deprivation** | | | |
| Index of Multiple Deprivation (England) | 13.83 (13.00) | 13.24 (12.49) | 0.0499 |
| **Outcome** | | | |
| Multimorbidity (≥2 diseases) | 111,262 (28.4%) | 26,640 (29.3%) | -0.0194 |

**Table S5. Feature importance (Scaled)**

| Feature | Gain | Split | SHAP | Mixed |
|---|---|---|---|---|
| Body mass index (BMI) | 0.840017 | 1 | 0.836649 | 0.878329 |
| Seen doctor (GP) for nerves, anxiety, tension or depression | 1 | 0.162423 | 1 | 0.790606 |
| Age | 0.370321 | 0.639157 | 0.61223 | 0.558485 |
| Systolic blood pressure | 0.11067 | 0.471466 | 0.37463 | 0.332849 |
| Hand grip strength | 0.108191 | 0.72432 | 0.2355 | 0.325878 |
| Frequency of tiredness/lethargy in last 2 weeks | 0.228761 | 0.263389 | 0.339145 | 0.29261 |
| Serious illness, injury, bereavement in the last 2 years | 0.190653 | 0.423178 | 0.2707 | 0.288808 |
| Sex | 0.043651 | 0.244074 | 0.404539 | 0.274201 |
| Unable to work because of sickness or disability | 0.454381 | 0.172081 | 0.182269 | 0.24775 |
| Seen psychiatrist for nerves, anxiety, tension or depression | 0.193315 | 0.17252 | 0.247116 | 0.215017 |
| Weight change compared with one year ago | 0.052017 | 0.250219 | 0.186633 | 0.168875 |
| Average total household income | 0.10628 | 0.157155 | 0.201078 | 0.166398 |
| Pulse rate | 0.041158 | 0.376646 | 0.108766 | 0.158834 |
| Sleeplessness/insomnia | 0.075241 | 0.132572 | 0.201124 | 0.152515 |
| In paid employment or self-employed | 0.138167 | 0.112818 | 0.138711 | 0.132102 |
| Alcohol intake frequency | 0.042268 | 0.208077 | 0.102973 | 0.114073 |
| Diastolic blood pressure | 0.037249 | 0.282704 | 0.056256 | 0.108116 |
| Suffer from nerves | 0.065572 | 0.115452 | 0.122479 | 0.106495 |
| Alcohol intake vs. 10 years previously | 0.035063 | 0.110184 | 0.134123 | 0.103373 |
| Getting up in the morning | 0.037039 | 0.190518 | 0.091008 | 0.102393 |
| Mood swings | 0.036356 | 0.084284 | 0.130638 | 0.095479 |
| Tens/highly strung | 0.031181 | 0.100966 | 0.110796 | 0.088435 |
| Sleep duration | 0.020267 | 0.242757 | 0.04016 | 0.085836 |
| Worrier/anxious feelings | 0.012413 | 0.089113 | 0.105381 | 0.078072 |
| Weight | 0.017487 | 0.227392 | 0.017347 | 0.069893 |
| Frequency of friends and family visits | 0.00926 | 0.103161 | 0.08279 | 0.0695 |
| Past tobacco smoking | 0.014158 | 0.114135 | 0.071628 | 0.067888 |
| Nap during the day | 0.017921 | 0.084284 | 0.076114 | 0.063608 |
| NVQ or HND or HNC or equivalent | 0.013203 | 0.11194 | 0.060663 | 0.061617 |
| MET minutes per week for walking | 0.018695 | 0.174276 | 0.016087 | 0.056286 |
| Retired | 0.028382 | 0.083845 | 0.054542 | 0.055328 |
| Other professional qualifications eg: nursing, teaching | 0.007723 | 0.105795 | 0.042766 | 0.049762 |

| Feature | Gain | Split | SHAP | Mixed |
|---|---|---|---|---|
| Hours of exposure to tobacco at home | 0.008685 | 0.124671 | 0.015724 | 0.041201 |
| MET minutes per week for vigorous activity | 0.00729 | 0.101844 | 0.02694 | 0.040753 |
| MET minutes per week for moderate activity | 0.008921 | 0.093942 | 0.029292 | 0.040362 |
| White | 0.006136 | 0.077261 | 0.025533 | 0.033616 |
| Fractured/broken bones in last 5 years | 0.006762 | 0.080773 | 0.015066 | 0.029417 |
| Number in household | 0.004864 | 0.072432 | 0.014497 | 0.026572 |
| Leisure/social activities | 0.004703 | 0.046093 | 0.025941 | 0.025669 |
| O levels/GCSEs or equivalent | 0.006977 | 0.090869 | 0.001194 | 0.025059 |
| Total number of qualifications reported | 0.006335 | 0.076822 | 0.007022 | 0.0243 |
| Frequency of depressed mood in last 2 weeks | 0.006816 | 0.053556 | 0.013129 | 0.021657 |
| Miserableness | 0.003604 | 0.036874 | 0.020679 | 0.020459 |
| Frequency of unenthusiasm/disinterest in last 2 weeks | 0.006806 | 0.058824 | 0.006749 | 0.019782 |
| Smoking status | 0.003889 | 0.044337 | 0.013346 | 0.018729 |
| Daytime dozing (narcolepsy) | 0.004695 | 0.050483 | 0.007172 | 0.017381 |
| Loneliness | 0.004445 | 0.037313 | 0.011221 | 0.01605 |
| A levels/AS levels or equivalent | 0.004555 | 0.057507 | 0.000157 | 0.015594 |
| Looking after home and/or family | 0.002219 | 0.03468 | 0.011939 | 0.015194 |
| Live with Husband, wife or partner | 0.001664 | 0.030729 | 0.013982 | 0.015089 |
| Risk taking | 0.00135 | 0.024583 | 0.016005 | 0.014486 |
| Worry too long after embarrassment | 0.001799 | 0.028534 | 0.009247 | 0.012207 |
| Able to confide | 0.001787 | 0.032924 | 0.006317 | 0.011836 |
| Frequency of tenseness/restlessness in last 2 weeks | 0.001746 | 0.026778 | 0.004225 | 0.009244 |
| Fed-up feelings | 0.002048 | 0.017559 | 0.00826 | 0.009032 |
| Number of vehicles in household | 0.001317 | 0.0259 | 0.001931 | 0.00777 |
| Unemployed | 0.001299 | 0.024144 | 0.001247 | 0.006984 |
| Alcohol drinker status | 0.001411 | 0.014486 | 0.004352 | 0.006151 |
| College or University degree | 0.001497 | 0.022827 | 4.06E-05 | 0.006101 |
| Above moderate/vigorous recommendation | 0.00095 | 0.010975 | 0.003752 | 0.004857 |
| Job involves heavy manual or physical work | 0.000929 | 0.014047 | 0.001945 | 0.004717 |
| Chronotype | 0.000765 | 0.012291 | 0.000977 | 0.003753 |
| IPAQ activity group | 0.000852 | 0.007902 | 0.00102 | 0.002699 |
| Guilty feeling | 0.000263 | 0.005707 | 0.002273 | 0.002629 |
| Live with son and/or daughter (include step-children) | 0.000476 | 0.007024 | 0.001107 | 0.002428 |

| Feature | Gain | Split | SHAP | Mixed |
|---|---|---|---|---|
| Smokers in household | 0.000295 | 0.006585 | 0.001132 | 0.002286 |
| Irritability | 0.000412 | 0.005707 | 0.001086 | 0.002073 |
| CSEs or equivalent | 0.0002 | 0.003951 | 0.000836 | 0.001456 |
| Asian or Asian British | 0.000285 | 0.003951 | 0.000452 | 0.001285 |
| Doing unpaid or voluntary work | 0.000205 | 0.003951 | 0.000176 | 0.001127 |
| Live with mother and/or father | 0.000205 | 0.003512 | 8.42E-05 | 0.000971 |
| Full or part-time student | 0.000132 | 0.003073 | 0.00014 | 0.000872 |
| Live with grandchild | 0.000211 | 0.003073 | 0.0001 | 0.000871 |
| Nervous feelings | 0.000201 | 0.002634 | 8.15E-05 | 0.000749 |
| Sensitivity/hurt feelings | 0.00012 | 0.002195 | 2.11E-05 | 0.000589 |
| Job involves walking or standing | 5.68E-05 | 0.001317 | 0.000179 | 0.000433 |
| Live with other unrelated | 5.24E-05 | 0.000878 | 3.52E-05 | 0.00025 |
| Live with brother and/or sister | 2.91E-05 | 0.000878 | 5.08E-06 | 0.000229 |
| Ever smoked | 2.13E-05 | 0.000439 | 0.000101 | 0.000165 |
| Mixed | 3.16E-05 | 0.000439 | 2.80E-06 | 0.000119 |
| Live with other related | 1.54E-05 | 0.000439 | 1.25E-06 | 0.000114 |
| Other ethnic group | 0 | 0 | 0 | 0 |
| Live with grandparent | 0 | 0 | 0 | 0 |
| Black or Black British | 0 | 0 | 0 | 0 |
| Chinese | 0 | 0 | 0 | 0 |

**Table S6. Diseases with a Prevalence Exceeding 1%**

| Disease | Number of Cases | Prevalence Rate |
|---|---|---|
| varicella [chickenpox] | 25810 | 5.149343 |
| zoster [herpes zoster] | 9485 | 1.892349 |
| measles | 18506 | 3.692125 |
| rubella [german measles] | 6607 | 1.31816 |
| viral warts | 11772 | 2.348627 |
| mumps | 12250 | 2.443993 |
| viral infection of unspecified site | 6091 | 1.215213 |
| dermatophytosis | 20955 | 4.180724 |
| other bacterial agents as the cause of diseases classified to other chapters | 5209 | 1.039246 |
| iron deficiency anaemia | 11215 | 2.2375 |
| other anaemias | 9699 | 1.935044 |
| other hypothyroidism | 26337 | 5.254484 |
| thyrotoxicosis [hyperthyroidism] | 5468 | 1.090919 |
| non-insulin-dependent diabetes mellitus | 13600 | 2.713331 |
| unspecified diabetes mellitus | 21884 | 4.366068 |
| obesity | 13524 | 2.698168 |
| disorders of lipoprotein metabolism and other lipidaemias | 75305 | 15.02407 |
| mental and behavioural disorders due to use of tobacco | 21975 | 4.384224 |
| depressive episode | 41246 | 8.228973 |
| other anxiety disorders | 18138 | 3.618705 |
| reaction to severe stress, and adjustment disorders | 10370 | 2.068915 |
| epilepsy | 5417 | 1.080744 |
| migraine | 21165 | 4.222621 |
| sleep disorders | 9560 | 1.907312 |
| mononeuropathies of upper limb | 13519 | 2.69717 |
| hordeolum and chalazion | 7718 | 1.539815 |
| other disorders of eyelid | 9731 | 1.941428 |
| disorders of lachrymal system | 6861 | 1.368835 |
| conjunctivitis | 17696 | 3.530522 |
| other cataract | 12763 | 2.546341 |
| glaucoma | 7640 | 1.524253 |
| visual disturbances | 5318 | 1.060992 |
| otitis externa | 12945 | 2.582652 |
| other disorders of external ear | 16026 | 3.197341 |
| suppurative and unspecified otitis media | 6821 | 1.360855 |
| disorders of vestibular function | 5768 | 1.150771 |
| other diseases of inner ear | 6798 | 1.356266 |
| other hearing loss | 5305 | 1.058398 |

| Disease | Number of Cases | Prevalence Rate |
|---|---|---|
| otalgia and effusion of ear | 10081 | 2.011256 |
| other disorders of ear, not elsewhere classified | 12318 | 2.457559 |
| essential (primary | 134417 | 26.81748 |
| angina pectoris | 20280 | 4.046055 |
| acute myocardial infarction | 11706 | 2.335459 |
| chronic ischaemic heart disease | 16209 | 3.233851 |
| atrial fibrillation and flutter | 8381 | 1.67209 |
| other cardiac arrhythmias | 5979 | 1.192868 |
| complications and ill-defined descriptions of heart disease | 7066 | 1.409735 |
| stroke, not specified as haemorrhage or infarction | 7032 | 1.402952 |
| other peripheral vascular diseases | 5609 | 1.119049 |
| phlebitis and thrombophlebitis | 12109 | 2.415862 |
| varicose veins of lower extremities | 15874 | 3.167015 |
| haemorrhoids | 14295 | 2.85199 |
| acute sinusitis | 18147 | 3.620501 |
| acute pharyngitis | 17865 | 3.564239 |
| acute tonsillitis | 18673 | 3.725443 |
| acute upper respiratory infections of multiple and unspecified sites | 31890 | 6.362361 |
| influenza, virus not identified | 7932 | 1.58251 |
| pneumonia, organism unspecified | 11891 | 2.372369 |
| unspecified acute lower respiratory infection | 31415 | 6.267594 |
| vasomotor and allergic rhinitis | 44157 | 8.809746 |
| chronic sinusitis | 9107 | 1.816934 |
| nasal polyp | 5287 | 1.054807 |
| other disorders of nose and nasal sinuses | 12110 | 2.416061 |
| bronchitis, not specified as acute or chronic | 8000 | 1.596077 |
| other chronic obstructive pulmonary disease | 9433 | 1.881974 |
| asthma | 59172 | 11.80538 |
| oesophagitis | 8243 | 1.644558 |
| gastro-oesophageal reflux disease | 36652 | 7.312426 |
| other diseases of oesophagus | 6650 | 1.326739 |
| gastric ulcer | 6613 | 1.319357 |
| duodenal ulcer | 6494 | 1.295615 |
| gastritis and duodenitis | 23149 | 4.618448 |
| dyspepsia | 10227 | 2.040385 |
| unspecified appendicitis | 7174 | 1.431282 |
| inguinal hernia | 15815 | 3.155244 |
| diaphragmatic hernia | 22978 | 4.584332 |
| other non-infective gastro-enteritis and colitis | 14321 | 2.857177 |

| Disease | Number of Cases | Prevalence Rate |
|---|---|---|
| diverticular disease of intestine | 16342 | 3.260386 |
| irritable bowel syndrome | 25794 | 5.146151 |
| other functional intestinal disorders | 8383 | 1.672489 |
| other diseases of anus and rectum | 21398 | 4.269107 |
| other diseases of intestine | 6091 | 1.215213 |
| haemorrhoids and perianal venous thrombosis | 21285 | 4.246562 |
| cholelithiasis | 16414 | 3.274751 |
| other diseases of digestive system | 6047 | 1.206435 |
| cutaneous abscess, furuncle and carbuncle | 7014 | 1.39936 |
| cellulitis | 14008 | 2.794731 |
| other local infections of skin and subcutaneous tissue | 7666 | 1.529441 |
| atopic dermatitis | 10180 | 2.031008 |
| seborrhoeic dermatitis | 6544 | 1.305591 |
| pruritus | 7076 | 1.41173 |
| other dermatitis | 34339 | 6.85096 |
| psoriasis | 11060 | 2.206576 |
| urticaria | 7493 | 1.494925 |
| skin changes due to chronic exposure to nonionising radiation | 7281 | 1.452629 |
| nail disorders | 6815 | 1.359658 |
| acne | 5723 | 1.141793 |
| follicular cysts of skin and subcutaneous tissue | 9379 | 1.871201 |
| seborrhoeic keratosis | 13842 | 2.761612 |
| other disorders of skin and subcutaneous tissue, not elsewhere classified | 23135 | 4.615655 |
| other rheumatoid arthritis | 6729 | 1.3425 |
| gout | 10631 | 2.120987 |
| other arthritis | 13244 | 2.642305 |
| coxarthrosis [arthrosis of hip] | 6886 | 1.373823 |
| gonarthrosis [arthrosis of knee] | 14214 | 2.83583 |
| other arthrosis | 52276 | 10.42956 |
| acquired deformities of fingers and toes | 11039 | 2.202387 |
| internal derangement of knee | 10850 | 2.164679 |
| other joint disorders, not elsewhere classified | 54406 | 10.85452 |
| spondylosis | 16998 | 3.391264 |
| other spondylopathies | 6347 | 1.266287 |
| other intervertebral disk disorders | 16145 | 3.221083 |
| dorsalgia | 47721 | 9.520798 |
| synovitis and tenosynovitis | 9792 | 1.953598 |
| other disorders of synovium and tendon | 6039 | 1.204839 |

| Disease | Number of Cases | Prevalence Rate |
|---|---|---|
| soft tissue disorders related to use, overuse and pressure | 8336 | 1.663112 |
| fibroblastic disorders | 11335 | 2.261441 |
| shoulder lesions | 16545 | 3.300886 |
| other enthesopathies | 22005 | 4.390209 |
| other soft tissue disorders, not elsewhere classified | 46115 | 9.200385 |
| osteoporosis without pathological fracture | 9868 | 1.968761 |
| chronic renal failure | 6043 | 1.205637 |
| calculus of kidney and ureter | 7049 | 1.406343 |
| cystitis | 11469 | 2.288176 |
| other disorders of bladder | 8484 | 1.692639 |
| other disorders of urinary system | 28313 | 5.648715 |
| hyperplasia of prostate | 13787 | 2.750639 |
| other disorders of male genital organs | 7399 | 1.476172 |
| benign mammary dysplasia | 10995 | 2.193608 |
| unspecified lump in breast | 13116 | 2.616768 |
| other disorders of breast | 9333 | 1.862023 |
| endometriosis | 8438 | 1.683462 |
| female genital prolapse | 11108 | 2.216153 |
| noninflammatory disorders of ovary, fallopian tube and broad ligament | 9728 | 1.940829 |
| polyp of female genital tract | 11994 | 2.392918 |
| dysplasia of cervix uteri | 7679 | 1.532034 |
| other noninflammatory disorders of vagina | 8677 | 1.731145 |
| excessive, frequent and irregular menstruation | 24723 | 4.932476 |
| other abnormal uterine and vaginal bleeding | 8486 | 1.693039 |
| pain and other conditions associated with female genital organs and menstrual cycle | 10508 | 2.096447 |
| menopausal and other perimenopausal disorders | 24589 | 4.905742 |
| spontaneous abortion | 5664 | 1.130022 |
| perineal laceration during delivery | 6765 | 1.349682 |
| single spontaneous delivery | 15203 | 3.033145 |

**Table S7. Diseases with a Prevalence Exceeding 5%**

| Disease | Number of Cases | Prevalence Rate |
|---|---|---|
| varicella [chickenpox] | 25810 | 5.149343 |
| other hypothyroidism | 26337 | 5.254484 |
| disorders of lipoprotein metabolism and other lipidaemias | 75305 | 15.02407 |
| depressive episode | 41246 | 8.228973 |
| essential (primary | 134417 | 26.81748 |
| acute upper respiratory infections of multiple and unspecified sites | 31890 | 6.362361 |
| unspecified acute lower respiratory infection | 31415 | 6.267594 |
| vasomotor and allergic rhinitis | 44157 | 8.809746 |
| asthma | 59172 | 11.80538 |
| gastro-oesophageal reflux disease | 36652 | 7.312426 |
| irritable bowel syndrome | 25794 | 5.146151 |
| other dermatitis | 34339 | 6.85096 |
| other arthrosis | 52276 | 10.42956 |
| other joint disorders, not elsewhere classified | 54406 | 10.85452 |
| dorsalgia | 47721 | 9.520798 |
| other soft tissue disorders, not elsewhere classified | 46115 | 9.200385 |
| other disorders of urinary system | 28313 | 5.648715 |

**Table S8. Genomic Risk Loci Summary**

| rsID | chr | pos | p | start | end | nSNPs | nGWASSNPs | nIndSigSNPs | IndSigSNPs | nLeadSNPs | LeadSNPs |
|---|---|---|---|---|---|---|---|---|---|---|---|
| rs1623927 | 1 | 1.08E+08 | 4.55E-08 | 1.08E+08 | 1.08E+08 | 47 | 9 | 1 | rs1623927 | 1 | rs1623927 |
| rs1077492 | 2 | 25286435 | 2.36E-10 | 25075281 | 25340610 | 171 | 37 | 3 | rs1077492; rs13387729; rs1530016 | 1 | rs1077492 |
| rs695238 | 3 | 49869158 | 6.79E-09 | 49734229 | 49960388 | 107 | 33 | 1 | rs695238 | 1 | rs695238 |
| rs1441433 | 4 | 1.02E+08 | 6.69E-13 | 1.02E+08 | 1.02E+08 | 320 | 43 | 2 | rs1441433; rs2583398 | 1 | rs1441433 |
| rs325521 | 5 | 1.04E+08 | 2.94E-10 | 1.04E+08 | 1.04E+08 | 174 | 25 | 1 | rs325521 | 1 | rs325521 |
| rs3996325 | 7 | 2053747 | 1.67E-08 | 1872921 | 2110850 | 78 | 10 | 1 | rs3996325 | 1 | rs3996325 |
| rs17701660 | 7 | 39419985 | 1.54E-08 | 39299374 | 39464732 | 49 | 9 | 1 | rs17701660 | 1 | rs17701660 |
| rs7849480 | 9 | 14648130 | 3.17E-08 | 14639666 | 14688114 | 46 | 8 | 1 | rs7849480 | 1 | rs7849480 |
| rs1421085 | 16 | 53800954 | 3.41E-10 | 53797908 | 53845487 | 112 | 16 | 1 | rs1421085 | 1 | rs1421085 |
| rs2229616 | 18 | 58039276 | 3.40E-08 | 57986923 | 58091681 | 56 | 1 | 1 | rs2229616 | 1 | rs2229616 |

**Table S9. Mapped Gene Summary**

| ensg | symbol | chr | IndSigSNPs |
|---|---|---|---|
| ENSG00000198890 | PRMT6 | 1 | rs1623927 |
| ENSG00000138031 | ADCY3 | 2 | rs13387729; rs1077492 |
| ENSG00000115137 | DNAJC27 | 2 | rs1077492; rs13387729 |
| ENSG00000084710 | EFR3B | 2 | rs1077492; rs1530016 |
| ENSG00000173531 | MST1 | 3 | rs695238 |
| ENSG00000164068 | RNF123 | 3 | rs695238 |
| ENSG00000176020 | AMIGO3 | 3 | rs695238 |
| ENSG00000173540 | GMPPB | 3 | rs695238 |
| ENSG00000176095 | IP6K1 | 3 | rs695238 |
| ENSG00000187492 | CDHR4 | 3 | rs695238 |
| ENSG00000185614 | FAM212A | 3 | rs695238 |
| ENSG00000182179 | UBA7 | 3 | rs695238 |
| ENSG00000183763 | TRAIP | 3 | rs695238 |
| ENSG00000164076 | CAMKV | 3 | rs695238 |
| ENSG00000164078 | MST1R | 3 | rs695238 |
| ENSG00000228008 | CTD-2330K9.3 | 3 | rs695238 |
| ENSG00000164077 | MON1A | 3 | rs695238 |
| ENSG00000138814 | PPP3CA | 4 | rs1441433; rs2583398 |
| ENSG00000254531 | AP001816.1 | 4 | rs2583398 |
| ENSG00000153064 | BANK1 | 4 | rs2583398 |
| ENSG00000002822 | MAD1L1 | 7 | rs3996325 |
| ENSG00000176349 | AC110781.3 | 7 | rs3996325 |

| ensg | symbol | chr | IndSigSNPs |
|---|---|---|---|
| ENSG00000106536 | POU6F2 | 7 | rs17701660 |
| ENSG00000175893 | ZDHHC21 | 9 | rs7849480 |
| ENSG00000140718 | FTO | 16 | rs1421085 |
| ENSG00000166603 | MC4R | 18 | rs2229616 |

**Table S10. Significant Genetic Correlations of RS4DV-Related Brain Regions**

| Brain Region | Beta | P-value (lr) | FDR-BH (lr) | rg | P-value (rg) | FDR-BH (rg) |
|---|---|---|---|---|---|---|
| Insular Cortex (left) | -0.00874 | 0.008668 | 0.019719 | -0.187 | 0.000563 | 0.019565 |
| Insular Cortex (right) | -0.01214 | 0.000222 | 0.00063 | -0.17 | 0.002918 | 0.048167 |
| Precentral Gyrus (right) | -0.00689 | 0.020384 | 0.042289 | -0.1611 | 0.003365 | 0.048167 |
| Paracingulate Gyrus (left) | 0.007862 | 0.008796 | 0.019719 | -0.1679 | 0.006881 | 0.048167 |
| Cingulate Gyrus, posterior division (left) | 0.014558 | 4.24E-06 | 1.64E-05 | -0.1945 | 0.004447 | 0.048167 |
| Temporal Fusiform Cortex, anterior division (left) | -0.01646 | 4.05E-10 | 3.13E-09 | -0.1926 | 0.001337 | 0.030978 |
| Temporal Fusiform Cortex, anterior division (right) | -0.01449 | 3.65E-08 | 2.12E-07 | -0.1958 | 0.006296 | 0.048167 |
| Amygdala (left) | -0.02114 | 3.54E-13 | 3.07E-12 | -0.2687 | 2.31E-05 | 0.003207 |
| Amygdala (right) | -0.02518 | 4.26E-17 | 5.31E-16 | -0.2047 | 0.002587 | 0.048167 |
| Ventral Striatum (right) | -0.01844 | 2.64E-12 | 2.16E-11 | -0.2001 | 0.000525 | 0.019565 |
| VIIIa Cerebellum (left) | -0.03587 | 2.59E-45 | 1.20E-43 | -0.1713 | 0.006781 | 0.048167 |
| VIIIa Cerebellum (right) | -0.03695 | 2.93E-49 | 4.07E-47 | -0.2349 | 0.004928 | 0.048167 |
| IX Cerebellum (right) | -0.0286 | 5.81E-34 | 1.61E-32 | -0.1796 | 0.00558 | 0.048167 |

### Supplementary Figures

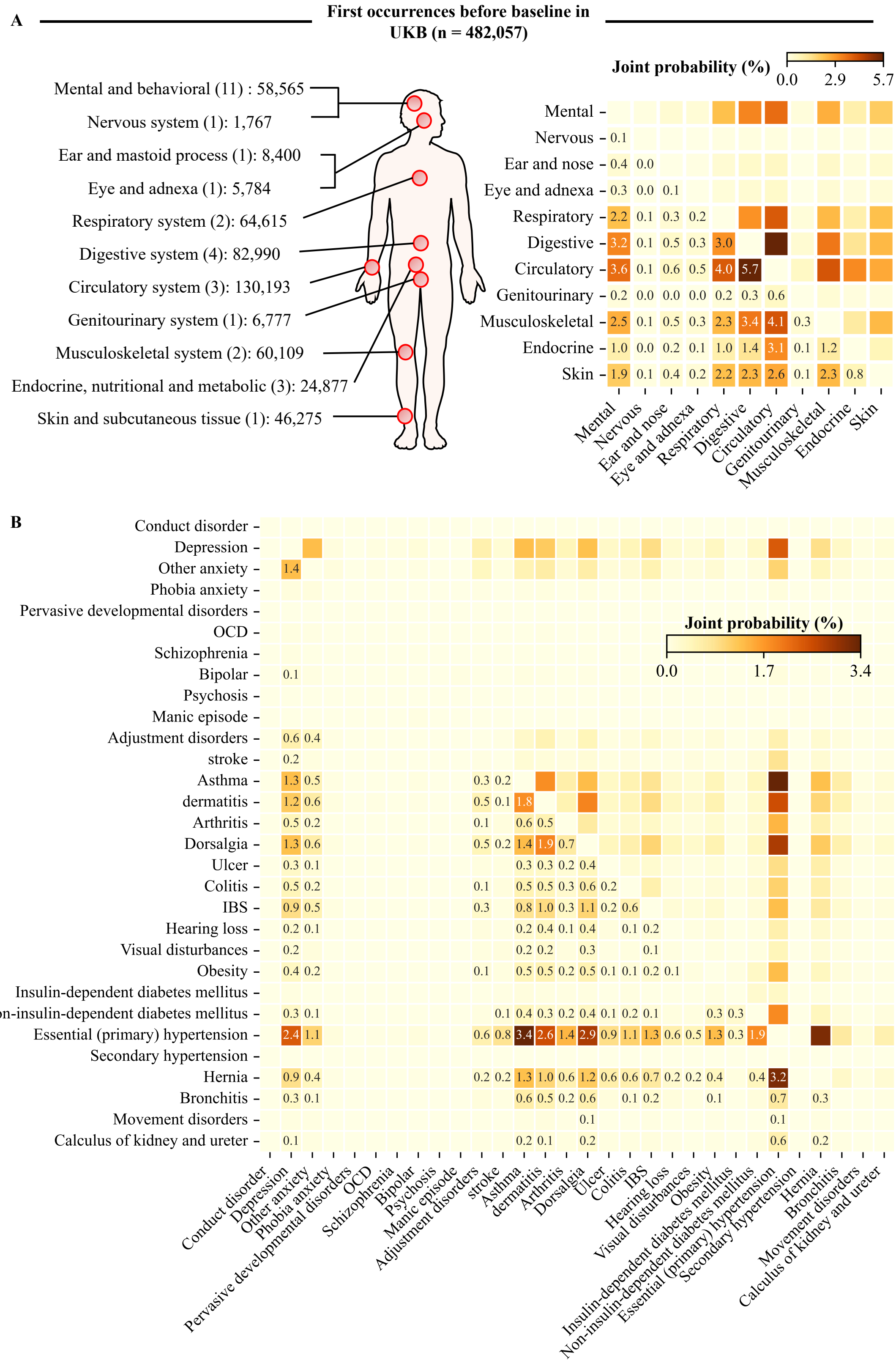

**Figure S1. Joint probability of co-occurrence between different disease at baseline.**

**(A)** Anatomical mapping of major disease categories and their case numbers based on first diagnosis records from 482,057 UKB participants, joint probability of co-occurrence between different disease categories at baseline. **(B)** Heatmap of joint probabilities (%) for co-occurrence of individual diseases. Each cell represents the proportion of individuals diagnosed with both diseases in the corresponding row and column. Darker shades indicate higher joint probabilities. The co-occurrence matrix highlights patterns of multimorbidity across both mental and physical health domains.

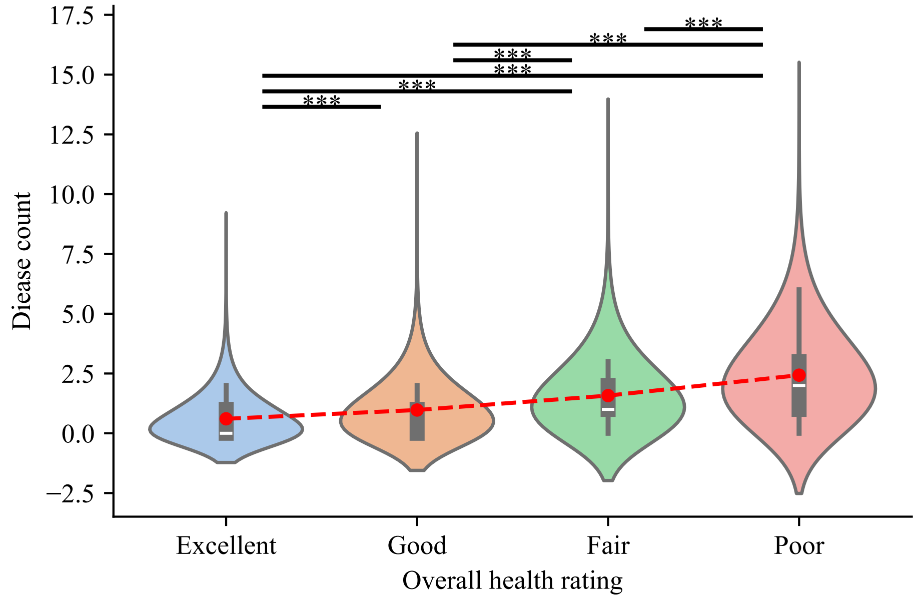

**Figure S2. Comparison of disease burden across self-rated health status groups.**

Violin plots illustrate the distribution of disease burden among four groups: Excellent, Good, Fair, and Poor health. The significance markers (*) denote $p$-values from two-sample t-tests between groups: *$p < 0.05$, **$p < 0.01$, ***$p < 0.001$.

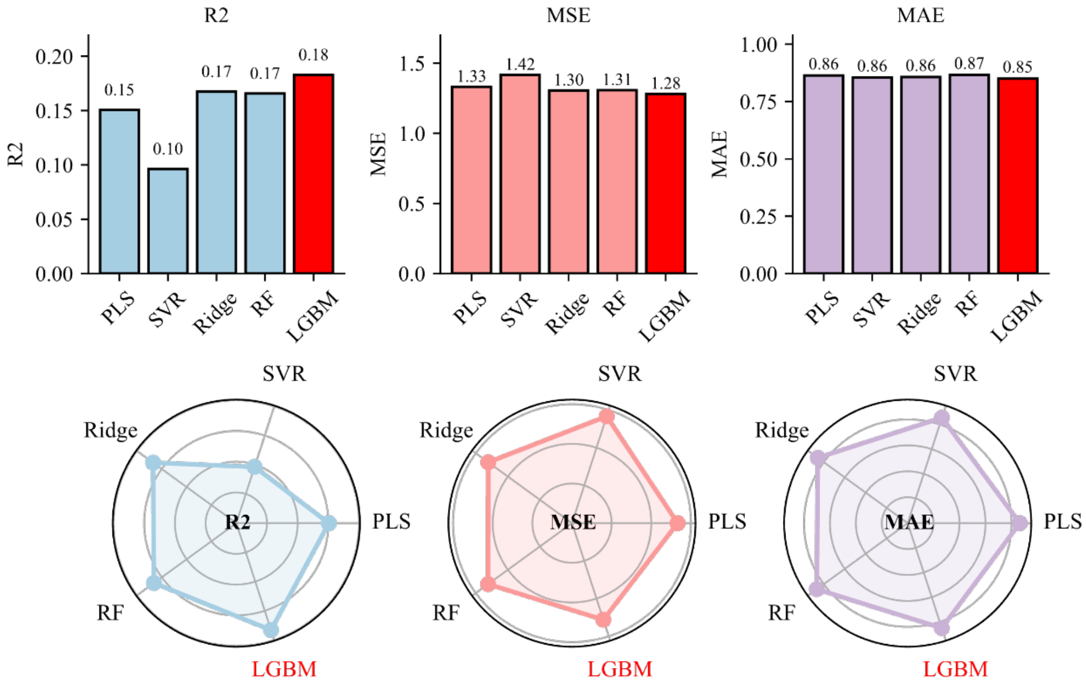

**Figure S3. Model performance.**

Comparison of model performance across five regression algorithms (LGBM, Ridge, PLS, SVR, and RF) using $R^2$, MSE, and MAE metrics in the independent test set. The bar and radar charts represent the relative performance of each model across different evaluation metrics. LGBM consistently showed superior performance, supporting its selection as the optimal model.

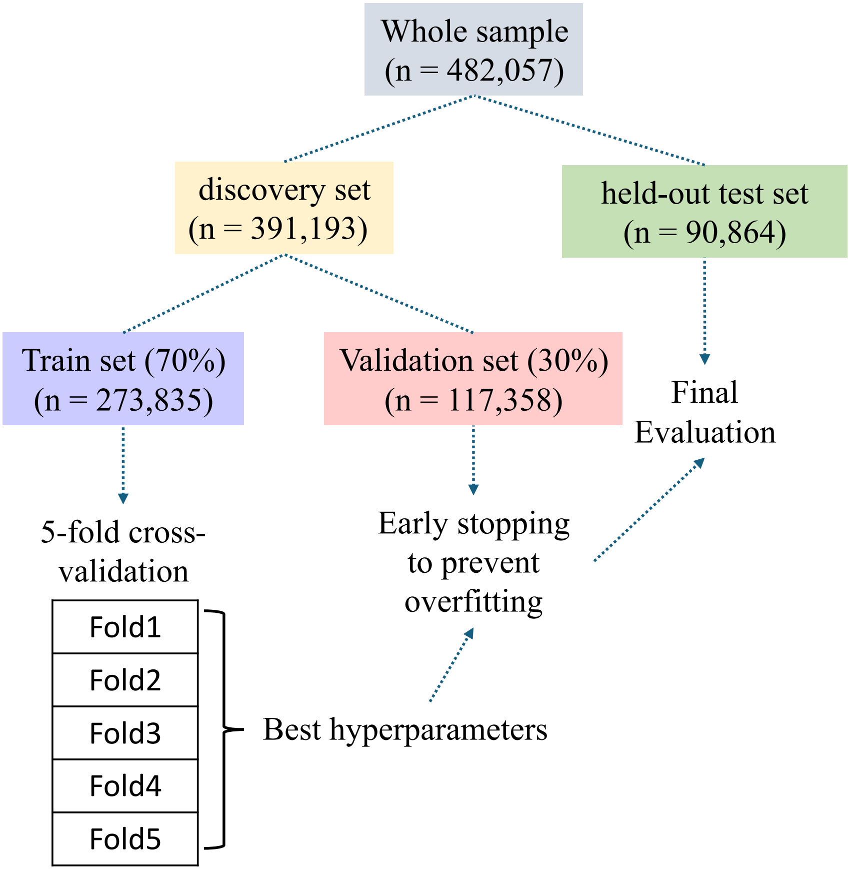

**Figure S4. Procedure for data partitioning.**

The initial total sample size (*n* = 482,057) was divided into the discovery set (*n* = 391,193) and the retained test set (*n* = 90,864). The discovery set is further divided into a training set (*n* = 273,835) and a validation set (*n* = 117,358) in a 7:3 ratio, which are respectively used for model training and early stopping to prevent overfitting.

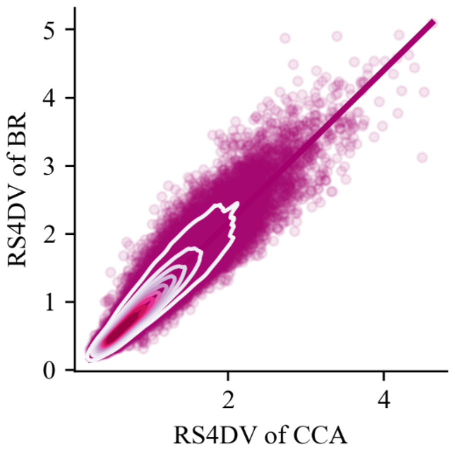

**Figure S5. Correlation between predictions from models trained on Bayesian-imputed data and complete-case analysis (CCA) data.**

Scatter plot showing the correlation between predictions derived from a model trained on Bayesian-imputed data ($n$ = 273,835) and those from a model trained on CCA data ($n$ = 20,248), both evaluated on an independent test set ($n$ = 90,864). Density contours represent the distribution of the predicted values. A high concordance was observed ($\rho = 0.95$, $p < 0.001$), demonstrating the robustness of the Bayesian imputation approach.

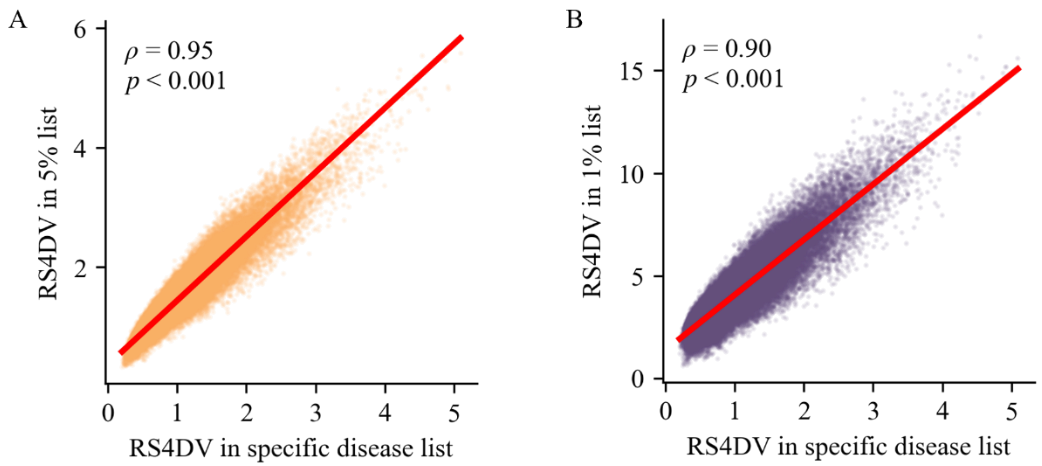

**Figure S6 Correlation between RS4DV derived from models trained on different disease lists.**

**(A)** Scatter plot showing the correlation between RS4DV derived from a model trained on ICD-10 diseases (Category 1712 of UKB) with a prevalence $\geqslant 5\%$ ($N = 17$), and those derived from a model trained on the specific disease list in the independent test set. **(B)** Scatter plot showing the correlation between RS4DV derived from a model trained on ICD-10 diseases with a prevalence $\geqslant 1\%$ ($N = 144$) in the independent test set, and those derived from a model trained on the specific disease list. Pearson correlation coefficient ($\rho$) is reported in each panel.

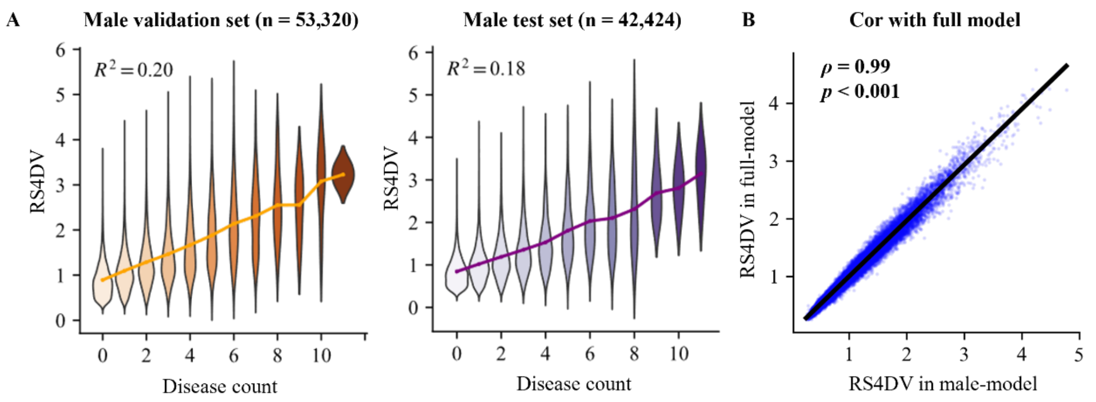

**Figure S7. Multivariable disease burden modeling based on the biopsychosocial framework (male-specific model).**

**(A)** Model performance in the male-only validation set ($n$ = 53,320) and independent test set ($n$ = 42,424). Violin plots show the predicted distribution of disease count using the final LGBM model, which achieved the best performance across five-fold cross-validation. (B) Scatter plot showing the Pearson correlation between predicted values from the male-specific model and those from the overall model in the independent test set.

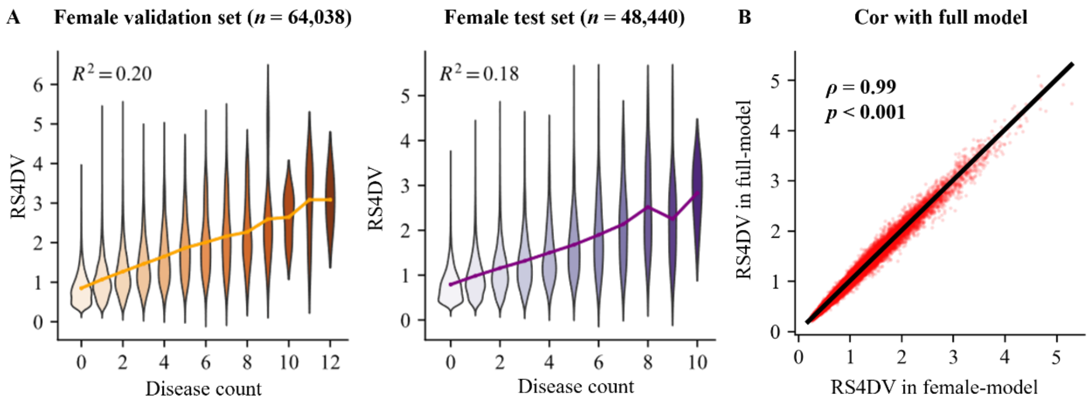

**Figure S8. Multivariable disease burden modeling based on the biopsychosocial framework (female-specific model).**

**(A)** Model performance in the female-only validation set (*n* = 64,038) and independent test set (*n* = 48,440). Violin plots show the predicted distribution of disease count using the final LGBM model, which achieved the best performance across five-fold cross-validation. **(B)** Scatter plot showing the correlation between predicted values from the female-specific model and those from the overall model in the independent test set.

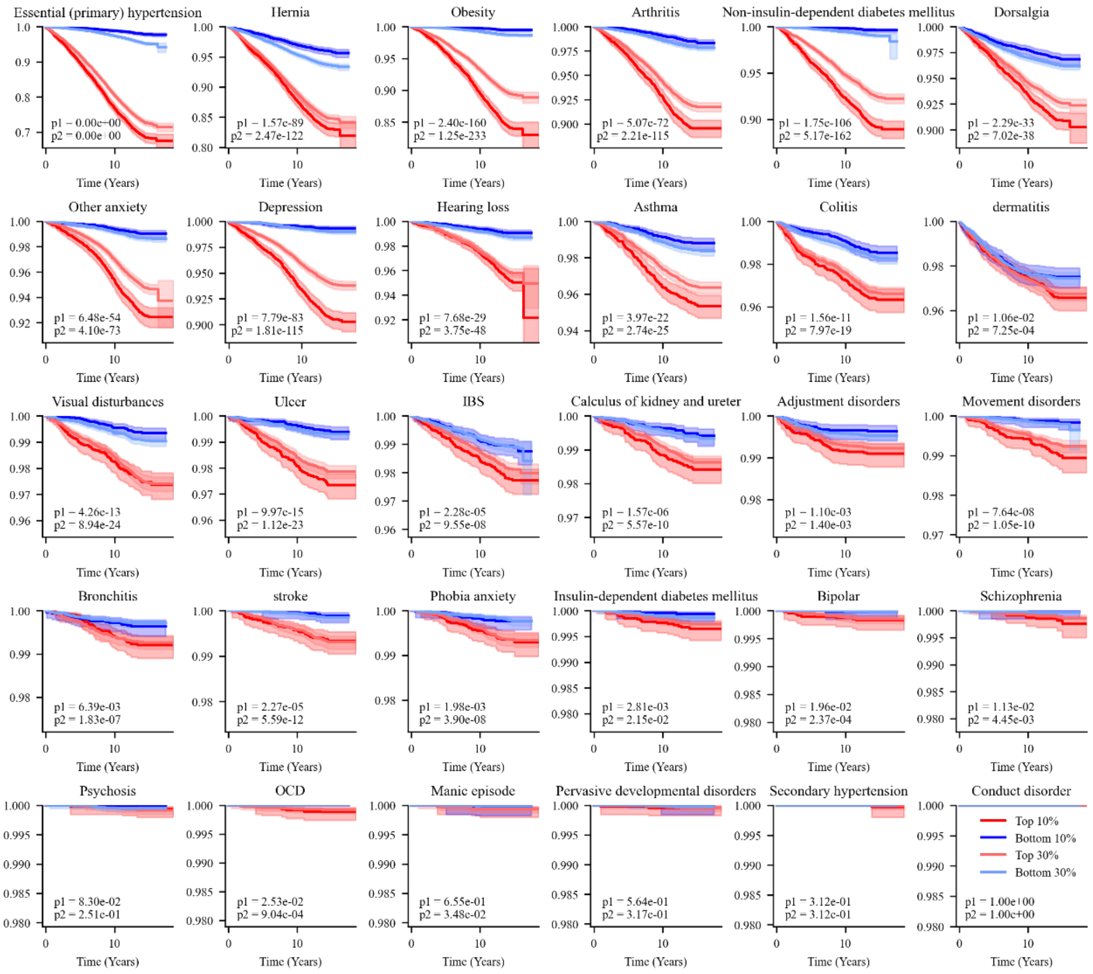

**Figure S9. Longitudinal assessment for individual disease of initially disease-free participants in validation subset.**

Kaplan-Meier survival curves compare the time to first incident disease between participants in the top 10%,30% (high risk) and bottom 10%,30% (low risk) of baseline RS4DV, average median follow-up time 14.7 years. The x-axis represents time since baseline (years), and the y-axis indicates the cumulative survival probability. *p*1: Top 10% VS Bottom 10 %, *p*2: Top 30% VS Bottom 30 %.

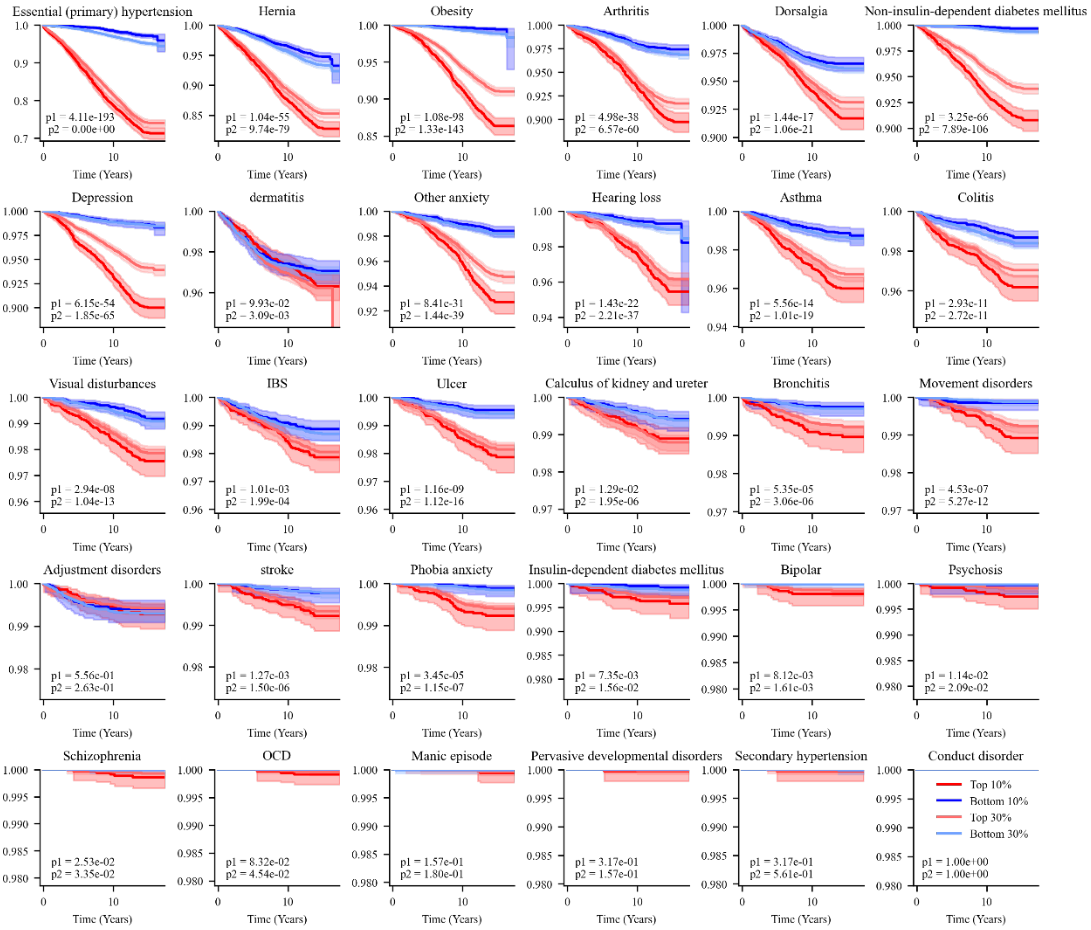

**Figure S10. Longitudinal assessment for individual disease of initially disease-free participants in test subset.**

Kaplan-Meier survival curves compare the time to first incident disease between participants in the top 10%,30% (high risk) and bottom 10%,30% (low risk) of baseline RS4DV, average median follow-up time 14.7 years. The x-axis represents time since baseline (years), and the y-axis indicates the cumulative survival probability. $p1$: Top 10% VS Bottom 10 %, $p2$: Top 30% VS Bottom 30 %.

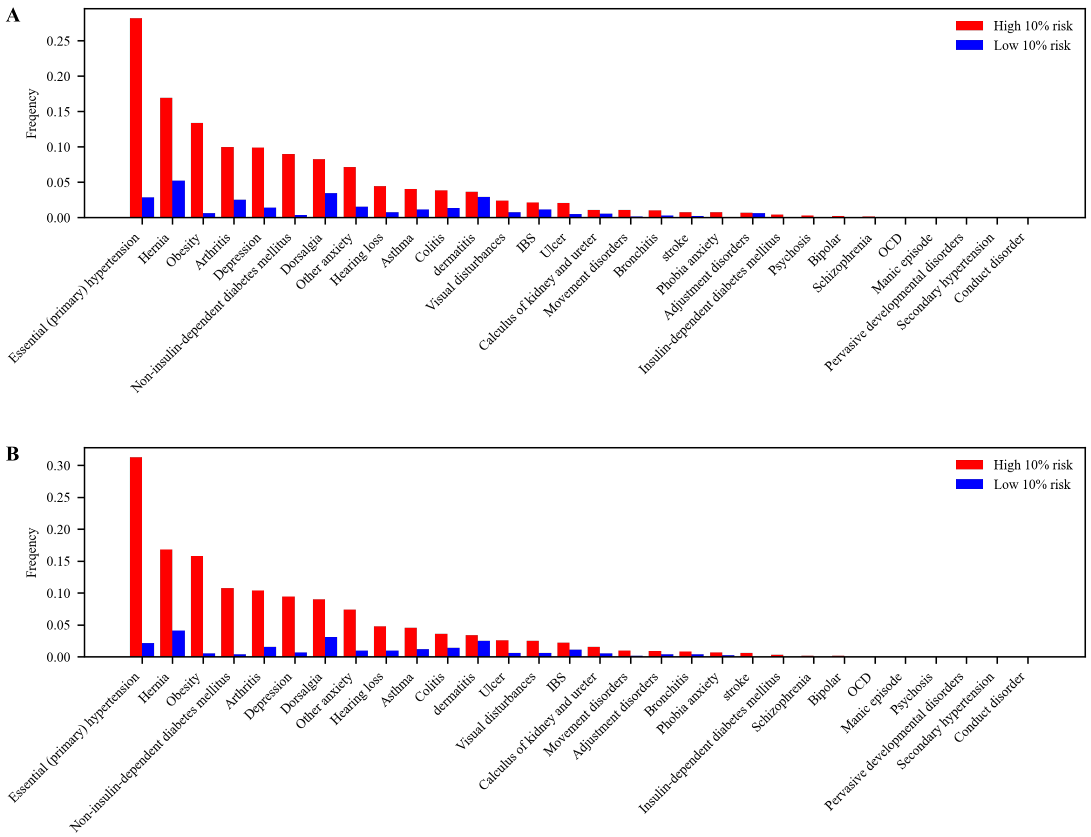

**Figure S11. Frequency of diseases between the high 10% risk group and the low 10% risk group during follow-up.**

**(A)** Bar chart illustrates the frequency distribution of different diseases in the validation subset, comparing the high 10% risk group (represented by red bars) and the low 10% risk group (represented by blue bars) over the follow-up period. **(B)** Bar chart illustrates the frequency distribution of different diseases in the test subset, comparing the high 10% risk group and the low 10% risk group over the follow-up period.

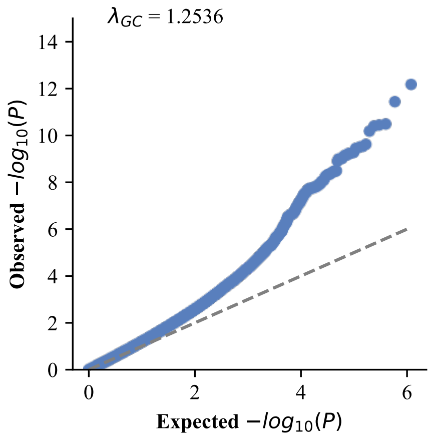

**Figure S12. QQ plot**

The QQ plot (λ = 1.25) demonstrates the expected versus observed *p* values, showing a modest inflation in association signals.

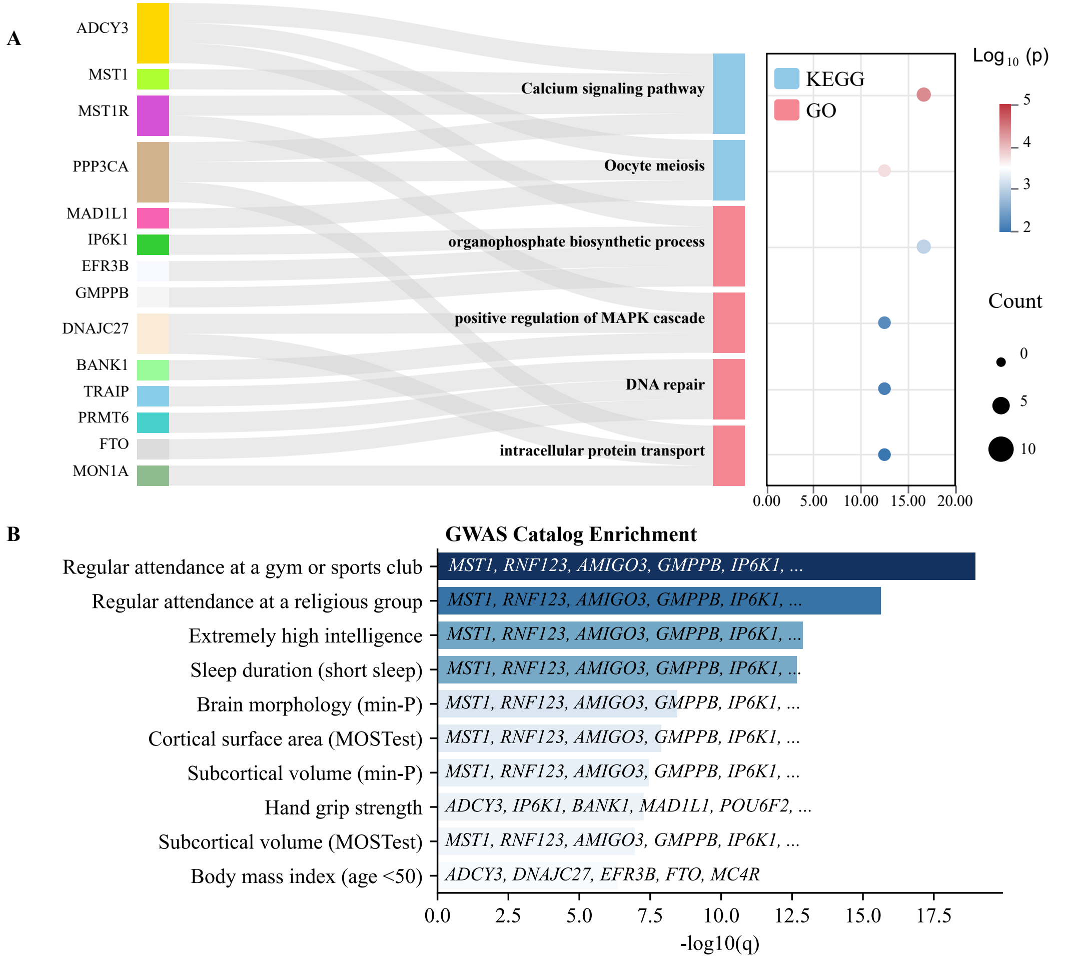

**Figure S13. Functional enrichment analysis of SNPs significantly associated with the RS4DV.**

**(A)** KEGG and Gene Ontology (GO) pathway enrichment analyses were performed on genes mapped from genome-wide significant SNPs. The results revealed biological pathways previously implicated in both physiological and pathological processes relevant to health and disease burden. **(B)** GWAS Catalog enrichment analysis identified significant over-representation of SNPs previously associated with complex traits.

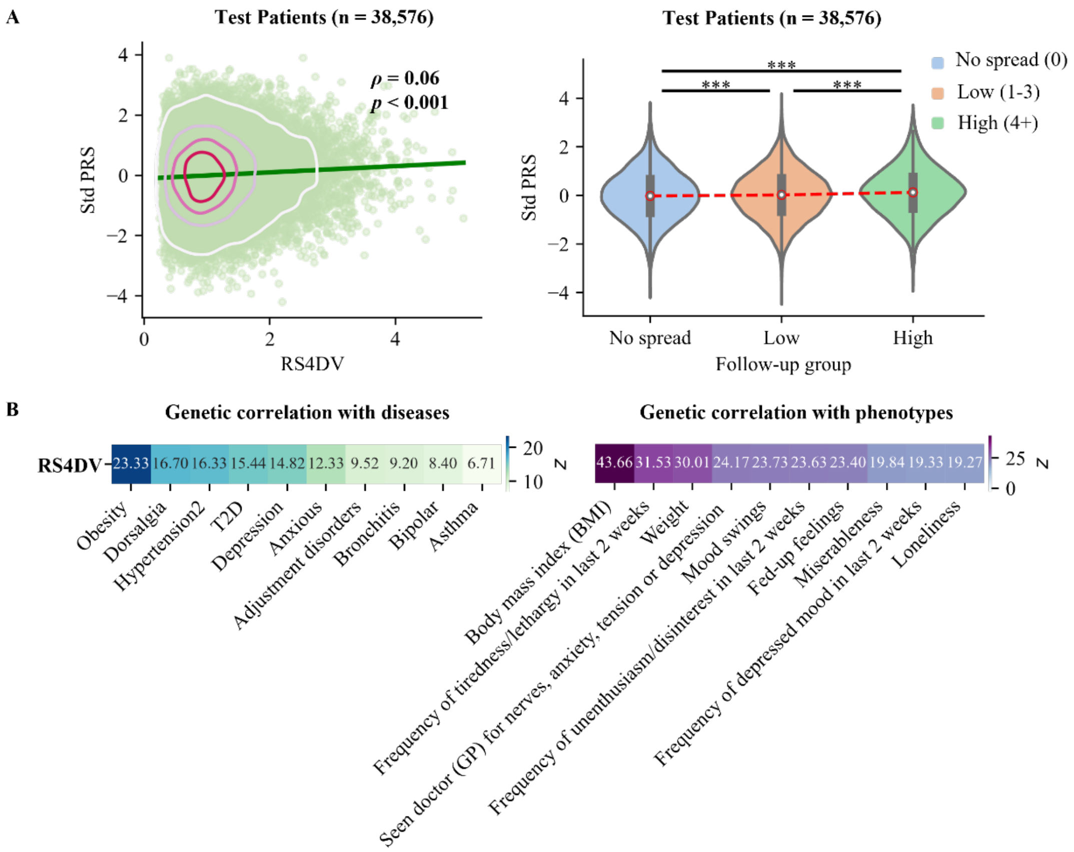

**Figure S14. Genetic correlations of RS4DV.**

**(A)** Scatter plot showing the association between the standardized polygenic risk score (PRS) and RS4DV among individuals with disease in the test patients set (Pearson $\rho = 0.06$, $p < 0.001$). Violin plots illustrate differences in standardized PRS across groups with no disease progression, low progression, and high progression. **(B)** Heatmap of genetic correlations between the RS4DV and selected diseases and phenotypic traits, showing the top 10 diseases and traits ranked by the absolute z-value of the genetic correlation estimates.

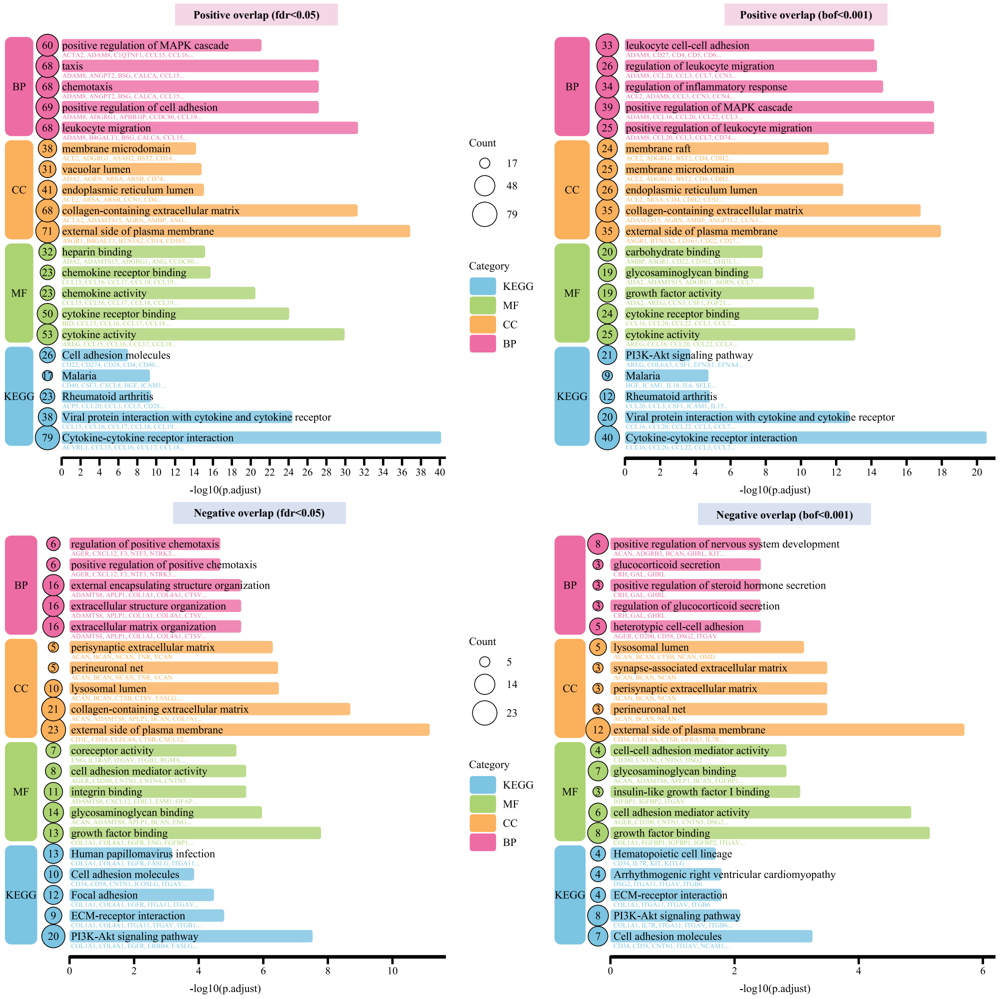

**Figure S15. Protein enrichment analysis.**

Functional enrichment analyses of differentially expressed proteins. The top-right panel shows significantly upregulated proteins after Bonferroni correction ($p < 0.001$), and the bottom-right panel shows significantly downregulated proteins under the same threshold. The top-left panel presents upregulated proteins identified using FDR correction ($p < 0.05$), and the bottom-left panel shows downregulated proteins identified with the same FDR threshold. Four enrichment analyses (GO-BP, GO-CC, GO-MF, and KEGG pathways) were conducted, with significance determined at FDR < 0.05.

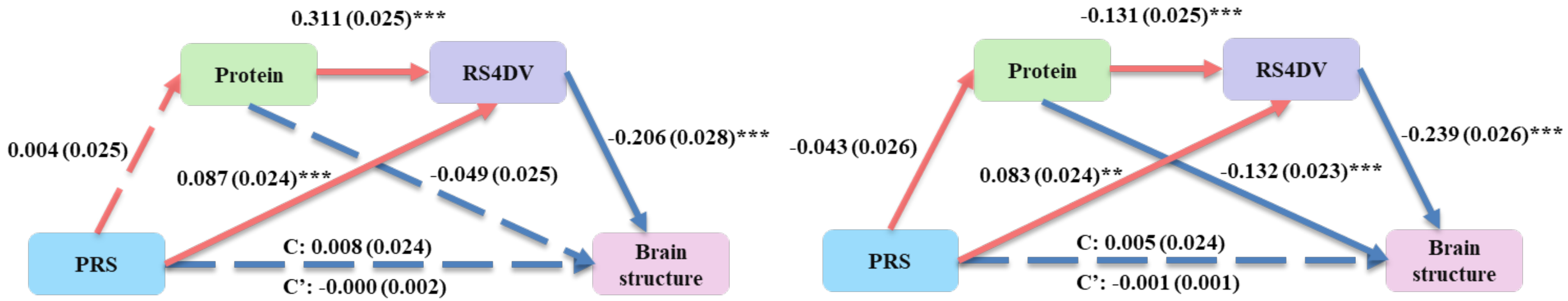

**Figure S16. Serial mediation model in diseases-free participants.**

Left: Serial mediation model (positive association protein model) linking PRS and brain structure in diseases-free participants ($n$ = 1,619). C' = direct effect of PRS on brain structure; C = total effect of PRS on brain structure. Right: Serial mediation model (negative association protein model) linking PRS and brain structure in disease participants. *$p < 0.05$, ***$p < 0.001$.

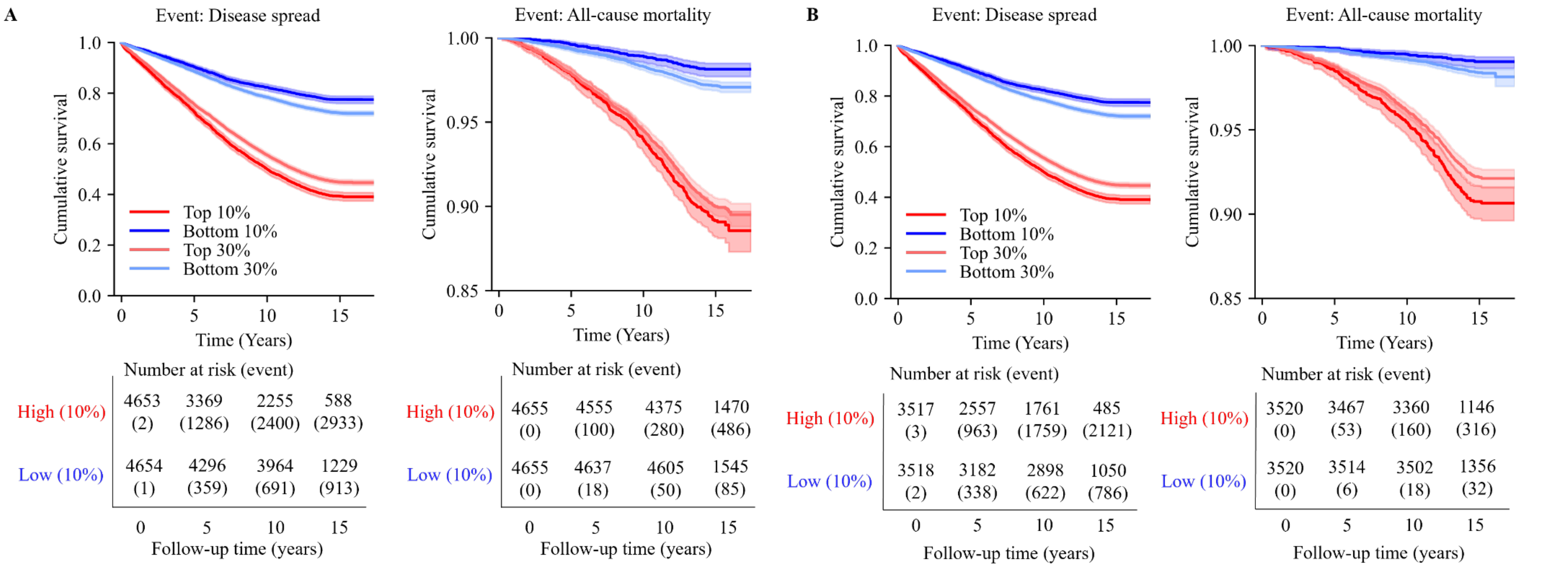

**Figure S17. 6-item RS4DV-based longitudinal assessment in disease-free participants**

**(A, B)** Kaplan-Meier survival curves illustrate the time-to-event analysis for participants classified into the top 10%, 30% (High Risk) and bottom 10%, 30% (Low Risk) of baseline 6-item RS4DV. Plots are shown for both the validation subset (left) and test subset (right) over average median follow-up time 14.7 years of follow-up. The x-axis denotes time since baseline (years), and the y-axis indicates cumulative survival probability. Numbers at risk (event) are provided below each curve to indicate how many individuals remain in the analysis at each time point.

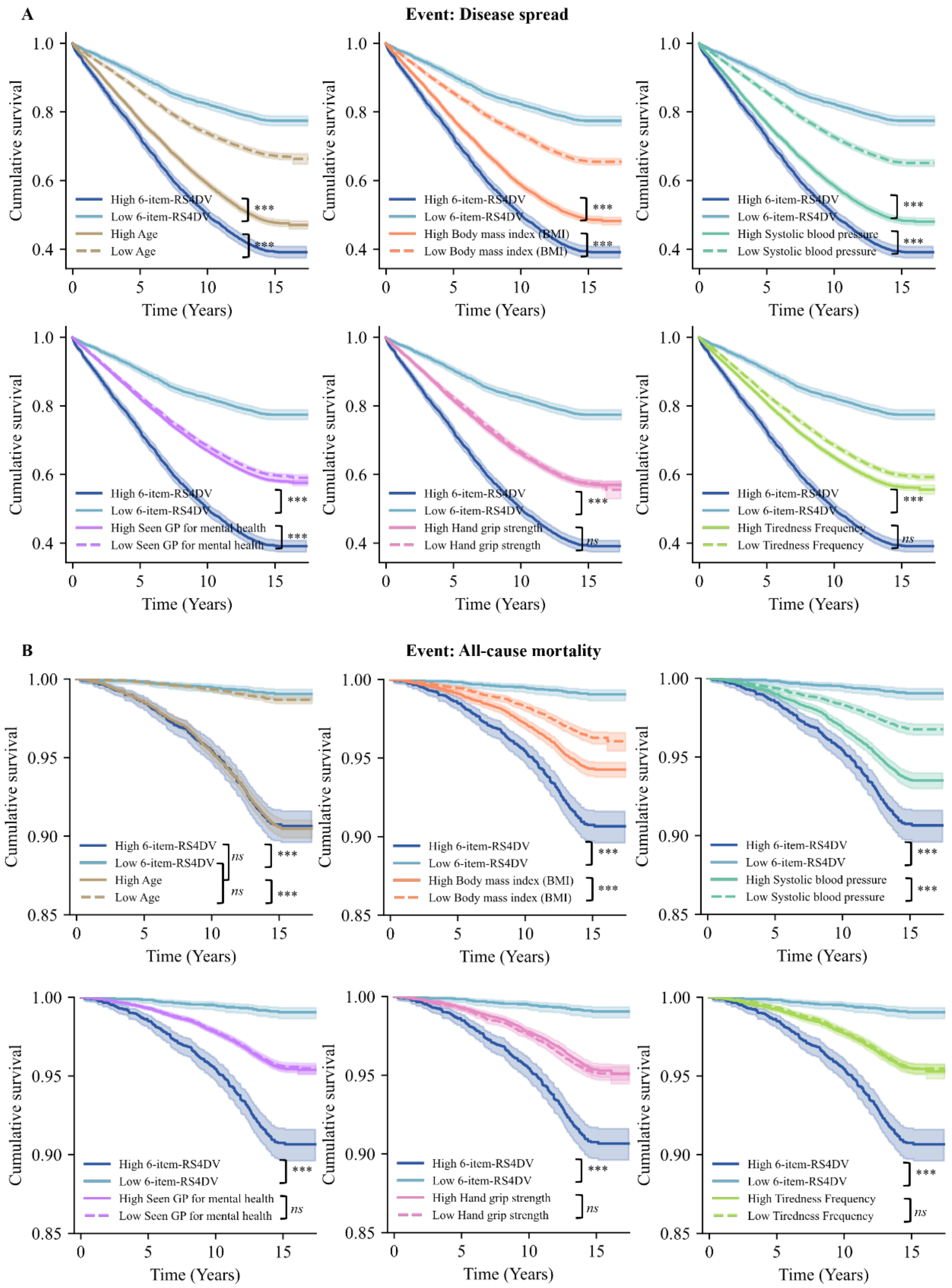

**Figure S18. Comparative survival analyses of the 6-item RS4DV and its constituent indicators in test subset disease-free participants.**

Kaplan-Meier survival curves comparing the prognostic performance of the composite 6-item RS4DV with each of its six original component indicators. Participants were stratified into high-risk and low-risk groups based on baseline values of the 6-item RS4DV or the corresponding individual indicator, as shown in each panel. Panel (A) presents time-to-event analyses for disease spread, while panel (B) presents time-to-event analyses for all-cause mortality. The x-axis denotes time since baseline (years), and the y-axis indicates cumulative survival probability. Shaded areas represent 95% confidence intervals. Curves are shown over a median follow-up period of 14.7 years. ***$p < 0.001$, ns $p > 0.05$.
